# A diagnostic decision-making protocol combines a new-generation of serological assay and PCR to fully resolve ambiguity in COVID-19 diagnosis

**DOI:** 10.1101/2020.08.11.20172452

**Authors:** Hu Cheng, Hao Chen, Yiting Li, Peiyan Zheng, Dayong Gu, Shiping He, Dongli Ma, Ruifang Wang, Jun Han, Zhongxin Lu, Xinyi Xia, Yi Deng, Lan Yang, Wenwen Xu, Shanhui Wu, Cuiying Liang, Hui Wang, Baoqing Sun, Nanshan Zhong, Hongwei Ma

## Abstract

The capacity to accurately diagnosis COVID-19 is essential for effective public health measures to manage the ongoing global pandemic, yet no presently available diagnostic technologies or clinical protocols can achieve full positive predictive value (PPV) and negative predictive value (NPV) performance. Two factors prevent accurate diagnosis: the failure of sampling methods (*e.g*., 40% false negatives from PCR testing of nasopharyngeal swabs) and sampling-time-dependent failures reflecting individual humoral responses of patients (e.g., serological testing outside of the sero-positive stage). Here, we report development of a diagnostic protocol that achieves full PPV and NPV based on a cohort of 500 confirmed COVID-19 cases, and present several discoveries about the sero-conversion dynamics throughout the disease course of COVID-19. The fundamental enabling technology for our study and diagnostic protocol—termed SANE, for Symptom (dpo)-Antibody-Nucleic acid-Epidemiological history—is our development of a peptide-protein hybrid microarray (PPHM) for COVID-19. The peptides comprising PPHMcovid-19 were selected based on clinical sample data, and give our technology the unique capacity to monitor a patient’s humoral response throughout the disease course. Among other assay-development related and clinically relevant findings, our use of PPHMcovid-19 revealed that 5% of COVID-19 patients are from an “early sero-reversion” subpopulation, thus explaining many of the mis-diagnoses we found in our comparative testing using PCR, CLIA, and PPHMcovid-19. Accordingly, the full SANE protocol incorporates orthogonal technologies to account for these patient variations, and successfully overcomes both the sampling method and sampling time limitations that have previously prevented doctors from achieving unambiguous, accurate diagnosis of COVID-19.

## Introduction

A second wave attack of severe acute respiratory syndrome coronavirus 2 (SARS-CoV-2) is expected, and the impact of the COVID-19 pandemic is predicted to last for decades (*1*). Therefore, there is an urgent need to review currently available diagnostic tools and use insights from such consideration to clarify the aims for new tool development (*2*). We believe that currently used diagnostic tools fall short in effectively containing the spread of the virus for at least two reasons: sampling-method- and sampling-time-related problems.

First, although used as the first choice and diagnostic gold standard, polymerase chain reaction (PCR)-based assays have unacceptably high rates of false negatives, *i.e*., approaching 40%, for detection of SARS-CoV-2 through non-invasive sampling methods such as nasopharyngeal swabs (*3, 4*). Non-invasive sampling has a strong potential to fail in the detection of silent spreaders (*i.e*., subjects who are asymptomatic or in the incubation period) of the virus. Specifically, negative PCR results can only be rigorously confirmed by invasive sampling, which is unlikely in asymptomatic subjects, given the current need for massive screening efforts at population scale. Such sampling-method-related problems cannot be improved through the detection technology, *i.e*., PCR technology itself.

Second, while it is the case that current serological assays have fewer problems than PCR-based methods from sampling-method-related artifacts, serological assays have stringent sampling time requirements that must fulfill well-defined criteria for successful confirmation of SARS-CoV-2 infection (*5, 6*). Consider that the period of immunoglobulin-G (IgG) production is typically very long for most patients, and these hosts remain sero-positive as they acquire immunity (*7, 8*). Thus, serological assays can capture information throughout the viral clearance period, when PCR-based assays would fail for accurate disease diagnosis by often yielding false negatives (*9*).

However, when sampling during the window period, serological assays can and do report false negative results (*5, 9*). These kinds of sampling-time-related problems can be partially improved through advancement of detection technologies. It should also be emphasized that serological assays have major issues connected to the selection and use of cut-off values: one must sacrifice sensitivity to achieve high specificity, or vice versa (*10, 11*). Thus, in present practice serological assays are generally only used to obtain sero-epidemiological data, rather than for screening in public health and clinical management (*12*).

As sampling-(method and time)-related problems cannot be fully resolved by diagnostic kits through technical means, achieving full sensitivity and specificity is not possible using currently available diagnostic kits; consequently, the need to conduct screening in low prevalence area remains unfulfilled (*13*). Recently, in a discovery phase trial using a small cohort of COVID-19 patients, we developed a peptide-protein hybrid microarray (PPHM) (*14*)that defines cut-off values in a fundamentally distinct manner from current serological assays such as lateral flow assays (*15*), enzyme-linked immunosorbent assays (ELISA) (16), and chemiluminescent immunoassays (CLIA) (11)(hereafter collectively referred to as “whole-protein-based assays”).

In the present study, we continue the development of this PPHM-based platform through the training and validation phases, using a large cohort of 500 COVID-19 cases. Our design for PPHM_covid-19_ exploits the potential of three aspects in enhancing the performance of COVID-19 diagnosis: i) a new way to select cut-off values; ii) differences in the IgG sero-dynamics of peptides vs. receptor binding domain (RBD) of spike protein; iii) a diagnostic decision-making protocol that enables full resolution of currently difficult diagnosis ambiguities in COVID-19 and that achieves full PPV and NPV performance; we term this protocol “symptom (dpo)-antibody-nucleic acid-epidemiological history” (SANE).

## Results

### PPHM defines cut-off values independent of the probe(s) and population(s) used in assay development

To empirically illustrate the challenges from sensitivity and specificity issues for selecting a suitable cut-off value for whole-protein-based assays, we used the SARS-CoV-2 RBD and N proteins in a microarray as discrete ELISA probes since our microarrays can be used in a full PPHM configuration or as discrete ELISA probes. The cut-off values were determined by first conducting tests in a training group that consisted of 333 control samples and 289 positive serum samples from PCR confirmed COVID-19 patients (sampled between March 6^th^ and 24^th^) (Fig. 1A, numbers in red and green, and Table S1); following typical practice, a signal value within the gray area was then selected as the cut-off value (Fig. 1C-F). The sensitivity and specificity were coupled through the cut-off value: that is, as the cut-off value was increased from 5 to 10 there was a decrease in the detection sensitivity of anti-RBD IgG from 85.8% to 85.1%, and a further increase in the cut-off value up to 20 dropped the sensitivity down to 82.0%. Moreover, these increases in cut-off value increased the specificity from 88.6% up to 99.4% (Fig. 1D).

**Fig. 1.**
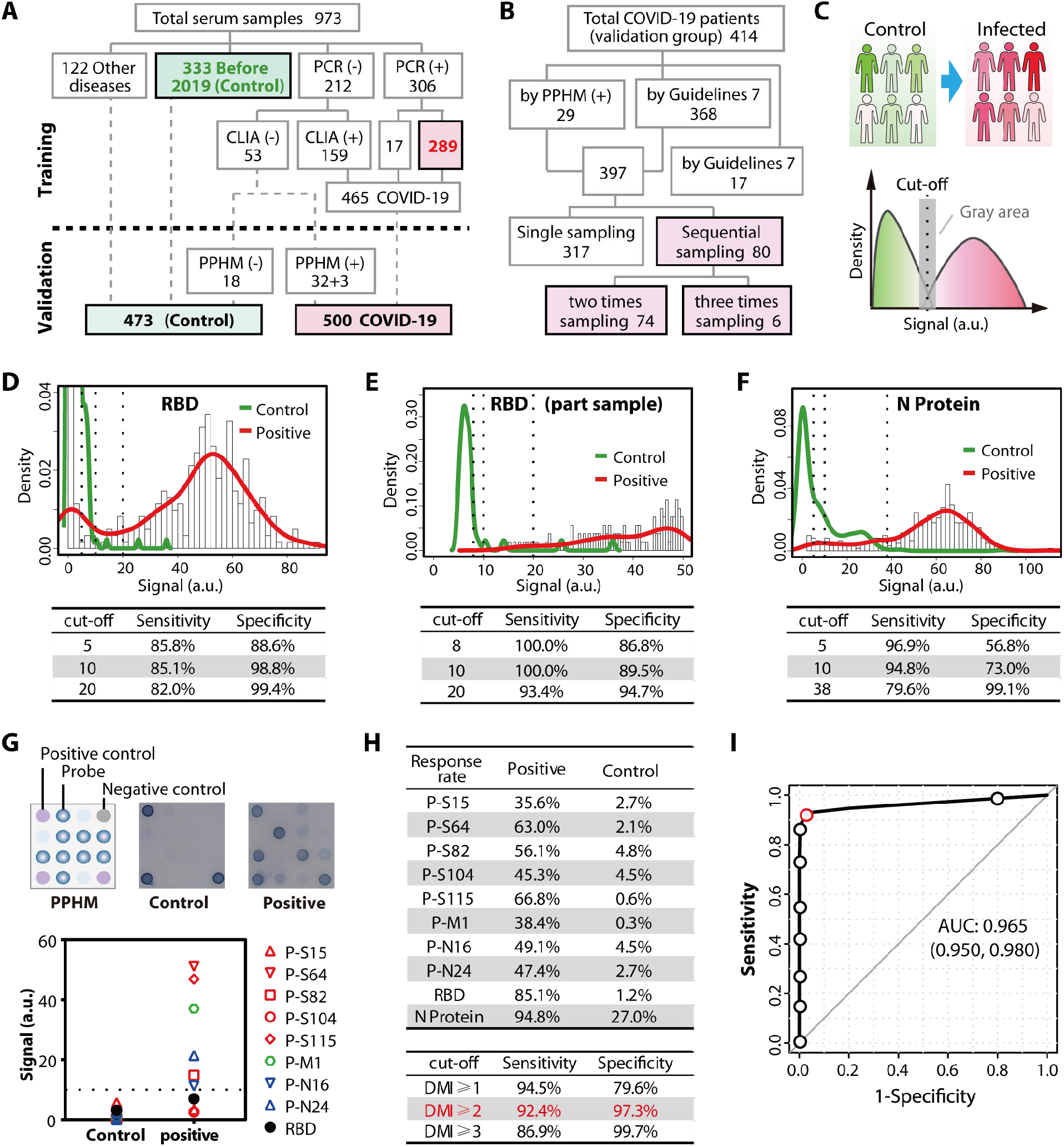
Impact of cut-off value and population on the sensitivity and specificity of whole-protein-based and PPHM assays. To demonstrate how determination of cut-off values can negatively impact sensitivity and specificity, we screened PPHMcovid-19 against a training group of 333 control and 289 COVID-19 positive serum samples. **(A)-(B)** Workflow for serum samples and patients. The initial disease status classification is performed prior to PPHMcovid-19. **(C)** Schematic illustration of whole-protein-based assays to determine cut-off values using the gray area at the overlapping interface of positive and negative test results. Since the microarray can be used in a full PPHM configuration or as discrete ELISA probes, we used RBD and N protein in a PPHM as discrete ELISA probes for the comparison between anti-RBD IgG signals for the training group **(D)** and for a patient subgroup **(E)** comprised of 38 control and 106 COVID-19 sera, which demonstrated that variation in the population used for assay development can obviously affect test performance evaluation outcomes. **(F)** Results for anti-(N protein) IgG. **(G)** Schematic illustration of the layout of PPHMcovid-19 (eight peptides as probes as well was the RBD protein) (top left), representative images of negative (top middle) and positive sera (top right), and signals of individual probes in PPHMcovid-19 for negative and positive serum samples (bottom). **(H)** Response rates for the N protein and for the probe molecules used in PPHM _covid-19_ (top). Sensitivity and specificity data for PPHMcovid-19 under different digital microarray index (DMI) values (*i.e*., different cut-off values) (bottom). **(I)** A receiver-operating-characteristic (ROC) curve of PPHMcovid-19. From right top to left bottom, DMI varies from 1 to 9, with the red circle indicating DMI = 2. The 0.965 AUC value indicated that PPHM is an outstanding serological test kit for COVID-19. The dashed lines indicate the cut-off values, including the arbitrary values selected by the assay developer, as well as the data-driven ones used for our PPHM assay.

Our data from these assays also emphasized that the cut-off value is heavily influenced by the particular population used for assay development. That is, for a serum subgroup of the training cohort that contained 106 COVID-19 and 38 control serum samples, anti-RBD IgG achieved 93.4% sensitivity and 94.7% specificity at a cut-off value of 20 (Fig. 1E). The absolute range of gray area and optimal cut-off value are also probe-dependent. For example, at a cut-off value of 10, anti-RBD IgG achieved 85.1% sensitivity and 98.8% specificity, while anti-(N protein) IgG achieved 94.8% sensitivity and 73.0% specificity (Fig. 1D, F and Fig. S1). Fundamentally, these results demonstrate that sensitivity and specificity are wholly dependent on cut-off value; and a practical implication of this fact is that assay developers must assess and explicitly account for population-specific impacts because we can now see that population characteristics obviously affect assay performance.

Seeking to resolve these cut-off value related issues affecting whole-protein-based assays, we developed a PPHM platform (Table S2) that defines cut-off values independent of i) the particular PPHM probes in the assay and ii) the population used in assay development. More specifically, PPHM employs two cut-off values: one at the individual probe level and the other at the whole microarray level. First, the cut-off value at the individual probe level in a PPHM is determined using a blank control. For example, when RBD (as a whole protein) was included as a probe for PPHMcovid-19, we detected response rates of 85.1% and 1.2% for COVID-19 and control samples, respectively (using a fixed and uniform cut-off value of 10) (Fig. 1G-H). Under this definition, a positive signal reveals the presence of IgGs in the tested serum that can recognize a PPHM probe, regardless of specificity (*17*).

For the second, whole-microarray-level cut-off value, we developed a Digital Microarray Index (DMI). This index represents the overall PPHM signal, and can be used to reliably determine if a tested serum sample is positive or negative for anti-(SARS-CoV-2) IgGs. Formally, DMI is the sum of the final probe values after positive and negative probe responses have been assigned values of 1 or 0, respectively.

The characteristics of the probes that were ultimately selected for inclusion in our PPHMcovid-19 included: high response rates for COVID-19 samples, low response rates for control samples, and complementary recognition to maximize coverage across the COVID-19 positive samples (Fig. 1H and Table S3). Our initial assessment of the PPHMcovid-19 was based on receiver-operating-characteristic (ROC) curves. The ability of PPHMcovid-19 to discriminate between COVID-19 and control serum samples was significant (*P* < 0.001), with an area under the curve (AUC, *i.e*., the overall accuracy of the test) equal to 0.965 (95% confidence interval, 0.95 to 0.98) (Fig. 1I). DMI = 2 was chosen as the optimal cut-off value for the PPHMcovid-19 because it provided the lowest false negative and false positive rates, achieving 97.3% specificity and 92.4% sensitivity for the training group.

We simulated the impact of population variation on the cut-off value of DMI by testing random patient subgroups sampled within the training group, and found that although the response rates for positive and negative serum samples changed, the performance of the PPHMcovid-19 assay remained consistent (Table S4). We thus obtained a PPHMcovid-19 comprising eight peptide probes and one whole protein-RBD probe, using a cut-off value of 10 for each individual probe and a cut-off value of 2 for DMI (*i.e*., the microarray level).

### Sampling time can confound the performance of serological assays

To demonstrate how sampling time can confound the performance of serological assays, we re-analyzed our PPHMcovid-19 results for the training group mentioned above, now focusing on differences in IgG sero-dynamics between the peptides and RBD. Owing to complex immunological factors (e.g., differences in immunological history, genetic background, and other contributing factors) and/or potential differences in infection conditions (e.g., viral load), i) patients vary in their incubation periods (*e.g*., the median incubation period for COVID-19 is four days (*18*) (Fig. 2A, bottom left); ii) patients experience differences in the duration for each of their respective sero-status stage (Fig. 2A, middle). Thus, even for samples collected at an identical dpo point, cohort sera collected at any given date will contain samples from all three sero-status stages, resulting in different performance of PPHMcovid-19 (Fig. S3).

**Fig. 2.**
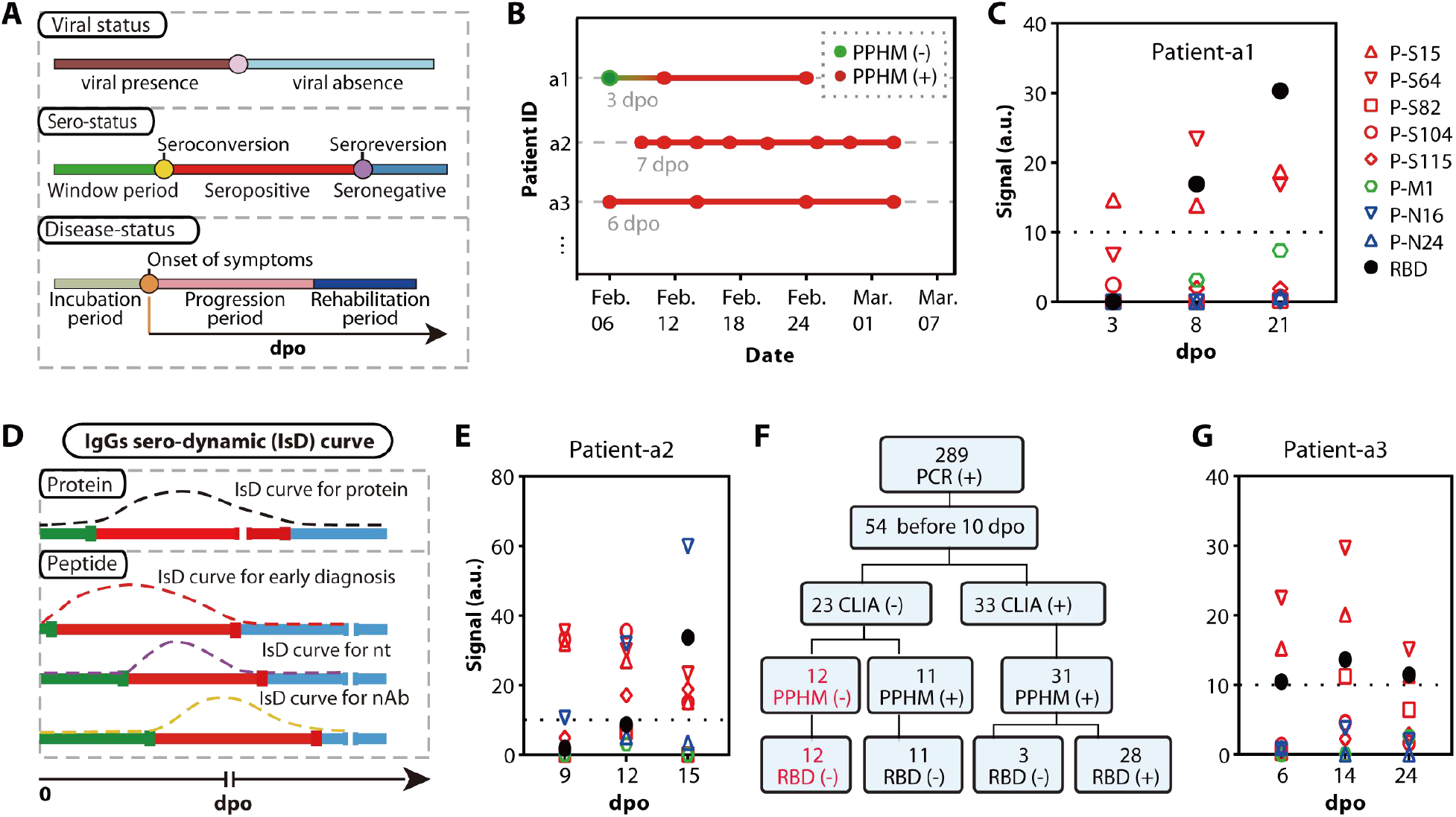
Well-known early sampling limitations can be overcome by PPHM. **(A)** Variation among hosts in the incubation period and throughout each sero-status stage precludes reliable confirmation of viral infection status in the sero-positive stage (red). That is, serum sampling can result in false negatives if a sample is collected during an unsuitable sero-status stage within the window period (green), especially for cross-sectional sera. **(B)** Representative clinically confirmed COVID-19 patients exhibiting the type of ambiguous results described in A. **(C)** Detailed signal values of nine PPHM probes for Patient-al from B. **(D)** Distinct IgG sero-dynamic (IsD) curves for peptides vs. whole protein data. At each day post onset (dpo), IsD for whole protein (top) yields only one signal during the entire sero-positive period, whereas PPHM has multiple IsD curves, therefore providing rich information about the duration of each sero-status period. Some peptide probes are detectable during the anti-whole protein IgG window period, and thus allow earlier detection of infection (middle). Some peptides can mirror PCR-based assays (second bottom), and others provide a direct measure of patient capacity for viral clearance (bottom). **(E)** Signals of each PPHM probe at different dpo for Patient-a2, an example case of early detection from D. **(F)** Workflow for 54 serum samples with early dpo according CLIA, PPHMcovid-19, and RBD (used as a discrete ELISA). **(G)** Patient-a3 is a representative of the highest frequency sample type, *i.e*., positive for COVID-19 at all three sampling points throughout the sero-positive period.

Using longitudinal serum samples for 3 patients that we obtained in another ongoing collaboration (Patients a1, a2, and a3), we were able to demonstrate the impact of sampling time on both viral- and sero-status. We found that although two confirmed COVID-19 patients were sampled at the same date (Feb. 6^th^), Patient-a1 had a false negative because of sampling within his window period (*i.e*., sampling too early) (Fig. 2B). In contrast, Patient-a2/a3 tested positive because of prolonged sero-positive periods. This example of a window period false negative result detected for Patient-a1 provides the opportunity to show how this problem of false negatives acquired in the window period (*i.e*., sampling too early) can be resolved: via continuous monitoring with PPHMcovid-19 (Fig. 2C). However, sequential sampling, especially starting from early dpo, is difficult to achieve in practice, especially when considering that whole-protein-based methods require the capture of a four-fold increase in the IgG level in a follow-up test (*11*). Below, we show how these sampling-time-related problems can be resolved by advancing diagnostic technologies.

### Some peptides enter into the sero-positive period while RBD is still in its window period

Notably, in the present study we found that some pathogen-derived peptides have potentially shorter window periods than whole proteins in the host, and thus allow for substantially earlier detection of infection (Fig. 2D, middle). For example, Patient-a2 showed positive test results at 9 dpo for three peptides (P-S15, P-S64, and P-S104), but did not show a positive result for RBD until 15 dpo, thus resulting in a positive diagnosis for COVID-19 by PPHM_covid-19_ at 9 dpo but a false negative diagnosis by CLIA (Fig. 2E).

To study the diagnosis capacity of PPHMcovid-19 in early dpo samples, we focused on 54 serum samples (dpo ≤ 10) of the 289 COVID-19 sera with positive PCR test results (Fig. 2F top). Of the 23 CLIA negative serum samples, ll(ll/54, 20.4%) were PPHMcovid-19 positive but RBD (used as a discrete ELSIA) negative (*i.e*., in agreement with CLIA test results), findings clearly underscoring the impressive early diagnosis ability of short peptides (Fig 2F bottom left). Of the 31 positive serum samples from the CLIA analysis, 3(3/54, 5.6%) were PPHM_covid-19_ positive but RBD negative (Fig 2F bottom right). It should be noted that the CLIA test results were all borderline positive (Table S5). Overall, the sensitivity achieved by PPHM _covid-19_ and CLIA for these 54 serum samples with early dpo was 42(42/54, 77.8%) and 31(31/54, 57.4%), respectively.

Note that we examined 12 serum samples from Figure 2F (in red) which were initially detected as PPHM _covid-19_ and RBD double negative in detail in Figures 3 and 4, below.

**Fig. 3.**
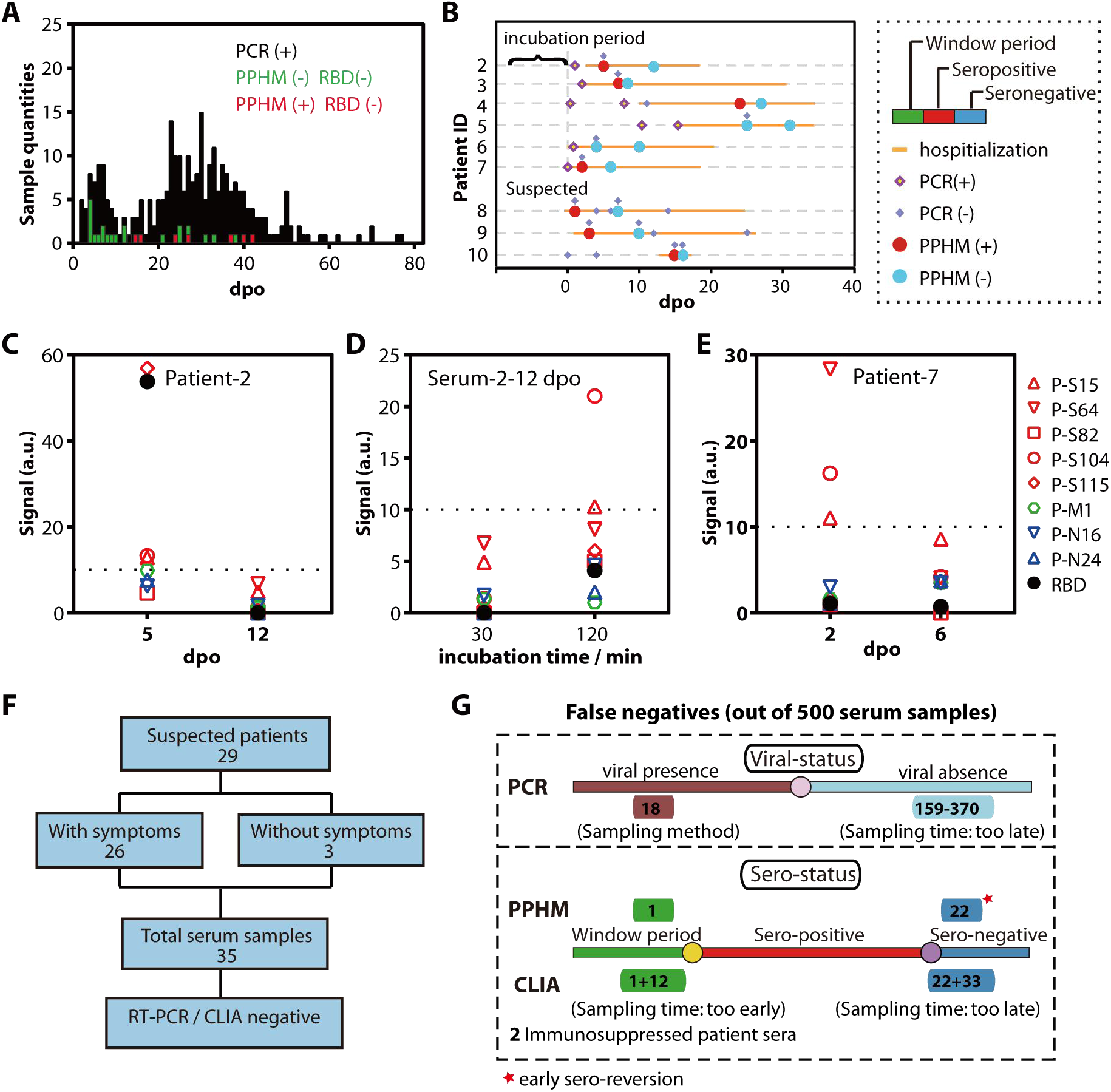
Unexpected late sampling limitations can be overcome by PPHMcovid-19. Distribution of 289 PCR positive serum samples of the training group against dpo; seven serum samples were PPHMCOVID-19 positive but RBD negative (red) and 22 serum samples were PPHMCOVID-19 negative (green). (**B**) PPHMCOVID-19 assay results and dpo together identified a sub-group with unusually fast antibody clearance (*i.e*., early sero-reversion). (**C**) Patient-2 underwent sero-reversion between 5 and 12 dpo. (D) When using the high-sensitivity iteration of PPHMCOVID-19, the detected status of Serum-2-12dpo (*i.e*., Serum-n(Patient-ID)-dpo) was converted from negative to positive, resulting in a DMI change from 0 to 2, *i.e*., positive of COVID-19. (**E**) Asymptomatic Patient-7 underwent sero-reversion between 2 and 6 dpo. The dashed line indicates cut-off value. (**F**) Workflow of 29 suspected patients, all the 35 serum samples were tested PPHMCOVID-19 positive. (**G**) All the false negative results of PCR, PPHM and CLIA caused by sampling method (*e.g*., non-invasive sampling) and sampling time (too early or late).

**Fig. 4.**
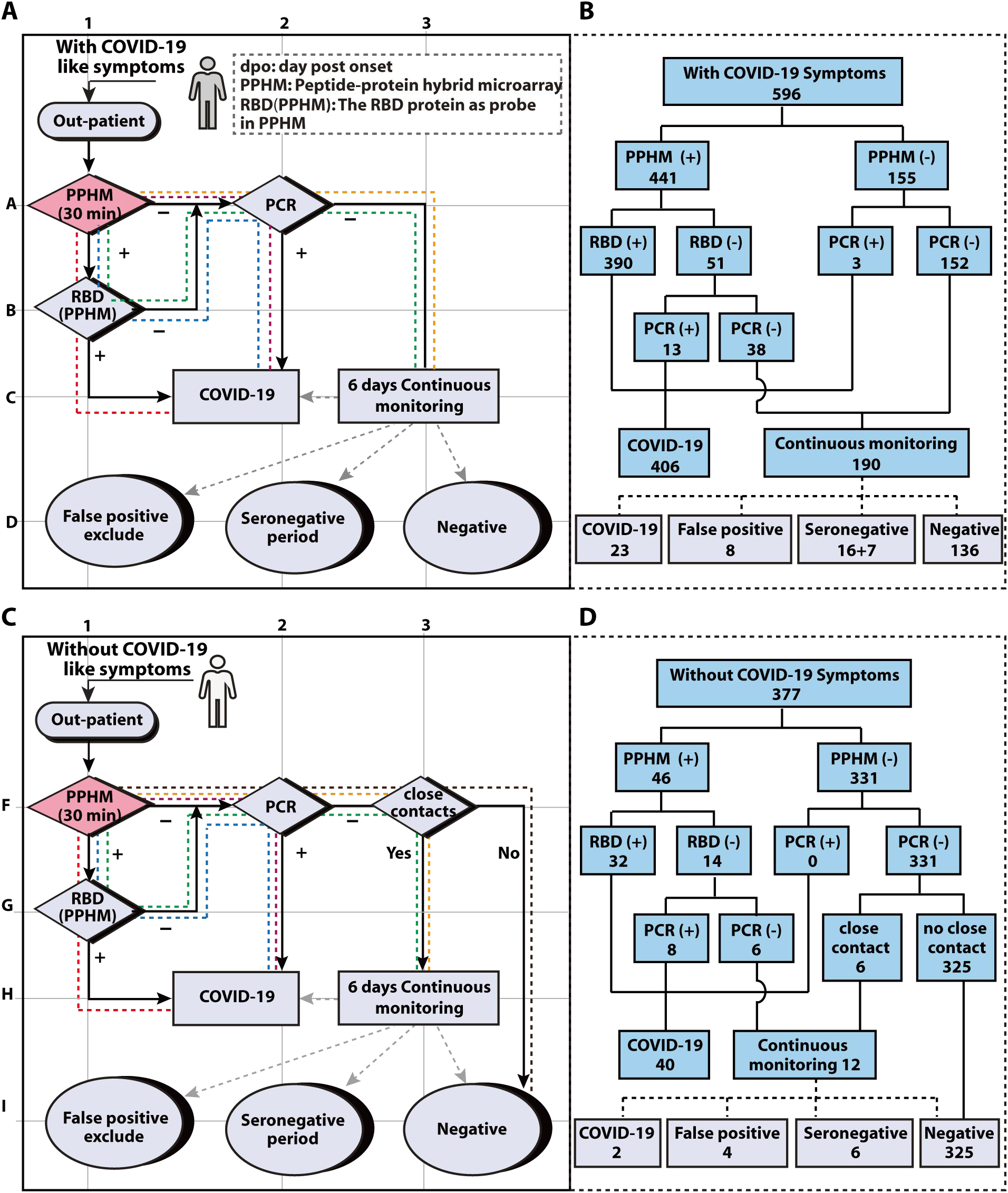
A proposed diagnostic decision-making protocol (SANE) for outpatients with and without COVID-19-like symptoms. Efficient clinical practice can be achieved by combining the merits of multiple assay types in diagnostic decision-making protocols (here, complete sensitivity and specificity for outpatients with COVID-19-like symptoms). (**A**) Patients with COVID-19-like symptoms are sorted and processed through one of five routes in sub-protocol 1. All start with a rapid iteration of PPHM_COVID-19_ assessment (grid A1) with subsequent PCR tests. The primary confirmation of SARS-CoV-2 infection is a positive result by either or both PPHM (both PPHMCOVID-19 and RBD) and PCR-based tests. Three routes (red, blue, and purple) identify positive samples (grid C2). The green and yellow routes accommodate samples which require continuous monitoring by rapid PPHMCOVID-19 assessment (for example in a quarantine context). (**B**) Workflow of outpatients. (**C**) Outpatients without COVID-19-like symptoms are assessed through one of six routes in sub-protocol 2. The flow is similar to sub-protocol 1 except, when both PPHMCOVID-19 and RBD tests are negative, the epidemiological history is used to determine whether a 6-day continuous monitoring (*i.e*., quarantine) is needed (grid C2) for these outpatients lacking obvious COVID-like-symptoms. (**D**) Workflow of outpatients.

### Some peptides remain in the sero-positive period even after RBD has entered into its sero-negative period

A clear trend from our data is that the cohort samples were largely obtained from the prolonged sero-positive period: of the 80 patients who contributed sequential serum samples, 71 patients had all positive PPHM _covid-19_ results (88.8%, 71/80) for serum samples collected from 1 to 77 dpo (Fig. 1B pink, Fig. S3).

Unexpectedly, we found that 7 serum samples (dpo >10) exhibited positive PPHMcovid-19 results while also yielding a negative RBD signal (Fig. 3A, red and Fig. S4). If these seven serum samples are not in the window period of RBD, such results indicate that pathogen-derived peptides apparently have a longer sero-positive period than RBD (Fig. S4). Moreover, PPHM _covid-19_ testing yielded a negative result for 22 serum samples (Fig. 3A, green). This unusual early sero-reversion was further supported by the results obtained using the high-sensitivity iteration of PPHMcovid-19, which can detect IgG levels below the limit of detection for the rapid iteration of PPHMcovid-19. For example, with rapid PPHMcovid-19, Serum-2-5dpo (*i.e*., Serum-n-dpo, where n is the Patient-ID) tested positive but Serum-2-12dpo tested negative (Fig. 3C). High sensitivity PPHMcovid-19 analysis of the Serum-2-12dpo detected anti-(P-S15) IgG and anti-(P-S104) IgG, indicating a recent sero-reversion event (Fig. 3D). The ability of high-sensitivity PPHMcovid-19 to discriminate positive and negative sera were validated in detail (Fig. 3C-E and Fig. S5). Thus, for analyzing these 29 (7+22) serum samples with early sero-reversion, CLIA detected none (0/29, 0%), whereas the sensitivity of PPHMcovid-19 was 24.1% (7/29).

### The 35 suspected subjects were all PPHMcovid-19 positive and confirmed as COVID-19 patients

To further explore the diagnosis capacity of PPHMcovid-19, we next examined the 35 suspected serum samples that were collected from 29 subjects, all of whom had an affirmative close contact history but were deemed negative based on both PCR and CLIA assays (Fig. 1A, bottom middle). Of these 29 subjects, 26 presented with COVID-19-like symptom whereas 3 did not (Fig 3F); however, these 3 asymptomatic subjects had radiology results indicating a positive COVID-19 status. Given their epidemiological histories and clinical features, now viewed in light of their positive PPHMcovid-19 results (Fig. S6), we can firmly conclude that all 29 of these subjects are indeed COVID-19 cases. There were 3 subjects who had one positive and one negative PPHMcovid-19 result, indicating early sero-reversion (Fig. 3B, Patient-8/9/10). We also noted that Serum-16-5dpo was in the sero-positive period for peptide-based analysis but was in the window period for RBD (Fig. S6); Serum-12-28dpo was in the sero-positive period for peptides but the sero-negative period for RBD. Collectively, these results clearly highlight that PPHMcovid-19 has overall higher sensitivity than whole-protein(RBD)-based assays (32/35 vs. 0/35). A total of 111 false negative PCR results was produced for these 29 patients, which could have been avoided if PPHMcovid-19 was used in the first place (Fig. S6). In total, our validation group comprised 500 serum samples (Fig. 1Abottom) from 414 COVID-19 patients (Fig. 1B top, and Table S1).

### PPHMcovid-19 can diagnose all four clinical classifications of COVID-19

Of the 500 serum samples from 414 COVID-19 patients, we conducted a layered analysis for detection sensitivity according to four clinical classifications of COVID-19 (*6*). For asymptomatic, moderate and severe classifications, PPHMcovid-19 showed superior performance over both whole-protein(RBD)- and PCR-based assays (Table 1 and Fig. S7); this superior performance was attributed to PPHM’s ability to assess COVID-19 status outside of the typical detection time frames used with whole-protein(RBD)- and PCR-based assays. The “critical classification” population was too small to enable any definitive conclusions about performance. However, we speculate that PCR-based assays may actually have an advantage for this classification: critical disease status is typically associated with viral-presence and is thus easy to sample (*i.e*., less prone to false negative due to non-invasive sampling). The relatively low sensitivity of PPHMcovid-19 for the asymptomatic classification can be attributed to the aforementioned “sampling too late” problem, especially for those patients exhibiting early sero-reversion. This is also complicated by the different methods that were used to determine the dpo for the asymptomatic patients (Fig. S8). Nevertheless, our data support that PPHMcovid-19 can be universally applied for all four classifications of COVID-19.

**Table 1.** Performance comparison among three diagnostic assays for all four clinical classifications of COVID-19.

| Classification | Total samples | PPHM |  | RBD<sup>a</sup> |  | PCR |  |
| --- | --- | --- | --- | --- | --- | --- | --- |
|  |  | Positive | Sensitivity | Positive | Sensitivity | Positive | Sensitivity |
| Asymptomatic | 48 | 42 | 87.5% | 31 | 64.6% | 34 | 70.8% |
| Moderate | 397 | 381 | 96.0% | 341 | 85.9% | 227 | 57.2% |
| Severe | 49 | 47 | 95.9% | 44 | 89.8% | 39 | 79.6% |
| Critical | 6 | 5 | 83.3% | 2 | 33.3% | 6 | 100.0% |
| Total | 500 | 475 | 95.0% | 418 | 83.6% | 306 | 61.2% |

<sup>a</sup> RBD protein in PPHM as discrete ELISA probe.

### Identification of a patient subgroup that exhibited early sero-reversion and is prone to false PPHMcovid-19 negative results

We also conducted a detailed study of 25 mis-diagnosed samples (*i.e*., false negative PPHMcovid-19 test results) from the 500 serum samples from all four clinical classifications of COVID-19, with a particular emphasis on patients having sequential serum samples. This analysis turned out to be an effective way of identifying low probability events. For example, Serum-7-2dpo showed a PPHMcovid-19 positive result by DMI (and was PCR positive), whereas Serum-7-6dpo was PPHMcovid-19 negative (and PCR negative) (Fig. 3B). We categorized Patient-7 into a patient subgroup characterized by early sero-reversion for the following reasons: i) medium to low anti-peptide IgG signal; ii) low DMI value; iii) short interval between viral-presence and absence, accompanied fast IgG elimination; iv) the basis used for dpo determination for asymptomatic patients (*i.e*., dpi could actually be much larger than dpo) (Fig. S8). Patient-8 from the asymptomatic class showed a similar trend, except that this subject was clinically confirmed as COVID-19 by a positive radiology result.

Serum-2-5dpo was PCR negative but PPHMcovid-19 positive (DMI = 4 and high RBD signal), indicating in the sero-positive period (Fig. 3C). Serum-2-12dpo tested PPHMcovid-19 negative (both DMI and RBD), indicating that IgG had been eliminated within 7 days after viral clearance (Fig. 3B-D). Thus, Patient-2 was also categorized into the early sero-reversion patient subgroup. Overall, we identified a total of 22 serum samples from 20(20/414, 4.8%) patients belonging to the early sero-reversion patient subgroup (Fig. 3B and Fig. S9). Note that a study of a larger early sero-reversion patient subgroup will be reported elsewhere.

This sub-group of patients characterized by early sero-reversion is apparently prone to false negative PPHMcovid-19 results; moreover, our detection of these patients motivated us to consider the testing-related-impacts of this and other types of low probability immunological events in considerable detail (see Discussion and Table S6). Very briefly, our theoretical analysis clearly indicated that low probability events will almost certainly have substantial problematic impacts on testing performed at the population level. Thus, we considered and accounted for the presence of such biological variation in the population as we developed our diagnostic decision-making protocol. That is, to avoid diagnosis failure caused by false negative or false positive PPHMcovid-19 results, the Symptom (dpo)-Antibody-Nucleic acid-Epidemiological history (SANE) protocol we developed incorporates orthogonal data sources in discrete steps to overcome the accuracy-sabotaging impacts from these improbable (yet certain-to-be-encountered) biological conditions in the population.

There were three patients with paired PCR and serum samples tested who were PCR positive but PPHMcovid-19 negative. One serum sample (Serum-172-7dpo) was in her window period and was identified as COVID-19 patient by positive PCR test result (Fig. S10). The other two serum samples were from COVID-19 patients in the critical classification. Follow-up analysis exploring one sample (Serum-38-21dpo) revealed that although the rapid iteration PPHMcovid-19 was negative, the high-sensitivity iteration PPHMcovid-19 result was positive (DMI = 2 but negative RBD signal) (Fig. 3G bottom and Fig. S10). Considering both the relatively late dpo and the patient’s condition (*i.e*., succumbing to late-stage stomach cancer 3 days), it is plausible that the negative rapid PPHMcovid-19 result can be understood as immunodeficiency. The identification of 2 immunosuppressed patients whose serum samples yielded negative results for both CLIA and PPHM but a positive result from a PCR test strongly demonstrate the need for diagnostic protocols that incorporate orthogonal detection technologies.

Overall, these efforts identified a significant number of false negative results from single-use diagnostic assays, including PCR, PPHMcovid-19, and CLIA. Out of the 500 positive serum samples, PCR produced 18 false negative results that were attributed to sampling method, and between 159 to 370 false negative results that were attributed to sampling time (too late) (Fig. 3G top). Among the two serological assays, CLIA produced 13 while PPHMcovid-19 produced only 1 false negative result due to sampling time (too early); CLIA produced 55 while PPHM _covid-19_ produced 22 false negative results due to sampling time (too late). Of the 55 false negative CLIA results due to sampling too late, 22 were false negatives from both PPHM _covid-19_ and CLIA, and 33 were PPHMcovid-19 positive but CLIA false negatives (Fig. 3G middle). This performance enhancement of PPHM _covid-19_ over CLIA is substantial; indeed, as we show in Figure 4, this improvement enabled our development of SANE protocol that incorporates PPHM _covid-19_ which has the potential to achieve full sensitivity and specificity for unambiguous COVID-19 diagnosis at the population level.

### A negative RBD signal in PPHMcovid-19 can be used to exclude false positive serum samples without close contact history

Among the 333 controls, we found that RBD showed a 98.8% specificity (cut-off value = 10) (Fig. 1D). Thus, we explored the use of an anti-RBD IgG negative result to help exclude PPHM _covid-19_ false positives. COVID-19 patients will become RBD positive during a 14-day quarantine period. For example, Patient-40 exhibited RBD sero-conversion between 6 and 7 dpo (Fig. S11). In contrast, subjects with false positive PPHMcovid-19 results (*i.e*., positive for peptides but RBD negative) will remain RBD negative during the self-isolation period. There were 12 false positives in the 473 control samples (Fig. 1A bottom and Fig. S12).

In practice, it is essential that test results accurately confirm or refute COVID-19. Although we demonstrated above that sampling time related problems can be partially resolved by advancing serological assay technologies, there remain PPHMcovid-19 false negative results, as well as false positive results (see Fig. 4). Thus, solutions beyond diagnostic technologies (including combined PPHM and PCR-based assays) such as diagnostic decision-making protocols that incorporate dpo/close contact history and other information are needed for efficient clinical practice, especially in cases of contradictory results. To visualize and apply these potential diagnostic outcomes in clinical practice, we developed a diagnostic decision-making protocol, *i.e*., Symptom-Antibody-Nucleic acid-Epidemiological history (SANE), including sub-protocol 1 (Fig. 4A) and sub-protocol 2 (Fig. 4B) for outpatients with and without COVID-19-like symptoms, respectively.

### Sub-protocol 1 of SANE for outpatients with COVID-19-like symptoms

Seeking to mimic the real context of clinical practice, we reconstructed our validation group for PPHMcovid-19 into a discovery cohort by assuming all serum samples for which we had four types of information (*i.e*., clinical symptoms, a PPHMcovid-19 test result, a PCR test result, and an epidemiological history) were from different subjects; arranging the data in this way yielded 596 and 377 serum samples with and without COVID-19-like symptoms, respectively (Fig. 4).

Sub-protocol 1 of SANE was applied to process the 596 serum samples with COVID-19-like symptoms. There are three routes that can directly identify outpatients as COVID-19 patients (Fig. 4A, coordinate C2): i) the red route gives 390 outpatients with PPHMcovid-19 and RBD positive results; ii) the blue route gives 13 outpatients with PPHMcovid-19 positive, RBD negative, and PCR positive results; iii) the purple route gives 3 outpatients with PPHMcovid-19 negative but PCR positive results (Fig. S10). The total confirmation of COVID-19 is 406.

Note that two routes require continuous monitoring to deliver definitive diagnosis results (Fig. 4A, coordinate C3): i) the green route (38 outpatients with PPHMcovid-19 positive, RBD and PCR negative results); and ii) the yellow route (152 outpatients with PPHMcovid-19, RBD and PCR all negative results). Thus, 190 outpatients in our study here require separate consideration because we did not have access to continuous monitoring data for them. Upon deployment of SANE, such patients would be assessed in their quarantine context based on the rapid iteration of PPHMcovid-19. Nevertheless, we were able to deduce from the data that we have that 23 outpatients should be understood as confirmed COVID-19: these 23 were identified when anti-RBD IgGs in the window period for the first sampling and then show PPHMcovid-19 and RBD positive results. We also deduced that eight outpatients can be understood as confirmed false positive: because they will remain RBD negative (Fig. S12). We strongly suspect that 16 (Fig. 3B and Fig. S9) and seven (Fig. S4) outpatients actually belong to respectively the early sero-reversion patient subgroup because of persistent negative PPHMcovid-19 and RBD test results. The remaining 136 outpatients represent confirmed negative COVID-19 cases, and very likely have other etiologies (Fig. S13).

## Sub-protocol 2 of SANE for subjects without COVID-19-like symptoms

Sub-protocol 2 of SANE was applied to the 377 serum samples from subjects without COVID-19-like symptoms. Similar to sub-protocol 1, there are three routes that can directly identify outpatients as asymptomatic COVID-19 patients (Fig. 4C, coordinate H2): these red, blue, and purple routes give 32, 8, and 0 COVID-19 patients, respectively. The total number of confirmed COVID-19 cases based on these samples is 40. Considering that these outpatients are without COVID-like symptoms, upon testing negative with PPHMcovid-19 and RBD, their epidemiological history is used to determine whether a 6-day continuous monitoring (*i.e*., self-isolation or quarantine) is needed (Fig. 4C, coordinate H2). If the outpatient has a close contact history, similar to sub-protocol 1, there are two routes that require continuous monitoring to delivery definitive diagnose results (Fig. 4C, coordinate H3). Note that the brown route gives 325 outpatients without close contact history as negative for COVID-19 and yellow route gives 6 outpatients with close contact history need to be continuously monitored (Fig. 4C, coordinate H3). As in our presentation of sub-protocol 1, our present data did not enable full implementation of SANE for sub-protocol 2; however, in theory a total of 12 outpatients would have been subjected to continuous monitoring by rapid iteration of PPHMcovid-19. We deduced that four of these are apparently false positive because they have no COVID-19 like symptoms and will remain RBD negative. The six serum samples that were previously confirmed from early sero-reversion patient subgroup will be diagnosed as negative because of persistent negative PPHMcovid-19 test results. Thus, the SANE protocol both overcomes the sampling-method-related problems with PCR-based assays and carefully solves the aforementioned problems related to sampling time (too early or late) (Fig. S13).

## Discussion

This study addressed a pressing issue that has to date limited the application of serological assays to clinical settings in the current fight against the global COVID-19 pandemic. Our study comprised two parts: the first being a validation of the PPHM platform (Fig. 1A) and the second being development of a diagnostic decision-making protocol (SANE) (Fig. 4). In addition to shining a light on some profound problems relating to the selection of cut-off values (see below), an emergent theme from the first part of our study is that currently deployed COVID-19 detection technologies are highly sensitive to factors which require correction. These include for example false negative samples resulting from sampling time issues (e.g., viral-absence, window period of IgG production, early sero-reversion, etc.) and sampling methods (e.g., non-invasive sampling) which do not actually reflect the known biology of SARS-CoV-2 replication. In the second part of our study, after establishment of PPHMcovid-19, SANE protocol was demonstrated as a method which can fully resolve disease-state ambiguities resulting from testing based on a single diagnostic technology.

Ideally, the causal relationship between the detection of anti-protein IgGs and virus infection ensures that only the infected population will show a positive response to pathogen-derived protein probes in serological tests. However, due to many known factors such as heterophilic antibodies (*19, 20*) and high concentration of mAb (*17*), this causal relationship can be masked by non-causal interactions (*i.e*., nonspecific interactions) so that a certain percentage of the non-infected population exhibit a positive anti-protein IgG signal. A cut-off value is thus needed to distinguish healthy and infected subjects (*13*). For example, in testing for SARS-CoV-2 infection, probe RBD showed 98.8% specificity whereas probe N protein showed only 73.0% specificity in this validation cohort (*5, 10, 16, 21*).

In contrast, PPHM defines cut-off values independent of probes in PPHM and the study population. The relationship between SARS-CoV-2 infection and anti-probe IgG response causally determines the probability of multiple probes being simultaneously positive, whereas negative results are determined by independent events (*i.e*., multiple non-specific interactions in one serum sample). Thus, we obtained a uniform cut-off value at 10 for individual probes and a universal cut-off value at 2 for DMI, as demonstrated in our previous papers for diagnosis of *P. falciparum* (*22*) and *Peste des petits ruminants* (*14*).

Our combined analysis of these multiple IsD curves motivated us to propose the following: that SARS-CoV-2 infection induces a humoral response characterized by i) anti-peptide IgGs appearing earlier than anti-RBD IgGs; ii) anti-peptide IgGs disappearing earlier than anti-RBD IgGs for most patients; iii) anti-peptide IgGs disappearing later than anti-RBD IgGs for a patient subgroup exhibiting early sero-reversion. Further, we suspect that the patient subgroup exhibiting early sero-reversion may be attributed to a “low dose” of SARS-CoV-2 infection. If so, perhaps this phenomenon could be detected in the low dose immunization cohort of various ongoing clinical trials for SARS-CoV-2 vaccine development (*7, 8*).

Despite the higher sensitivity and specificity available through serological testing as demonstrated in this paper and others (*10, 23*), PCR-based assays remain the most widely used diagnostic method for several reasons. For example, regardless of the exceptionally high false negative rate, false positives are exceedingly rare in PCR-based assays (*9*). Furthermore, false negative rates can be expected to be much lower in screening of the general population because the prevalence rate of SARS-CoV-2 is low in the general population. In contrast, serological tests may lead to both false negative and false positive results.

While potentially leading to epidemic viral spread (*24*), a high rate of false negative PCR results may be psychologically easier to manage in public health as compared to even a minimal number false positives, because they can potentially lead to unnecessary panic among the general population; these factors have resulted in the wide adoption of a less-reliable diagnostic method (*25*). Given the urgent need for diagnostic methods that provide 100% PPV or NPV, we hypothesized that improved serological assays could be used initially and then confirmed by PCR-based assays for COVID-19 diagnosis and screening of general populations through validation even in low prevalence areas.

Pursuing this, we designed and tested SANE protocol, to comprehensively address long-standing issues that plague serological testing and testing for SARS-CoV-2 infection, in particular. i) False negatives acquired by PCR-based assays; ii) false negatives generated in serological tests; and iii) false positives observed in serological test results. In this SANE protocol, PPHM is used first and supplemented by PCR because PPHM alone provides the highest sensitivity of all detection methods examined here (Table 1). Furthermore, our work also supports that the success of SANE depends on PPHMcovid-19 (Fig. S11). Highly notably, and suggesting the clinical and public health utility of our SANE protocol, performance assessments revealed that SANE achieves full PPV and NPV. We are unaware of any other COVID-19 testing protocols that achieve this fundamentally enabling level of diagnostic performance, and submit that SANE therefore warrants further examination for application in screening of the general population, even in low prevalence areas. Our ongoing work involves testing of 3000 sera from 3000 patients, and we are working with a variety of clinical stakeholders to deploy PPHMcovid-19 technology and SANE in both biomedical research and public health.

There are several limitations in this study, mainly due to two unexpected discoveries: the fact that peptides may enter into the sero-positive period while RBD is still in its window period, and the aforementioned patient subgroup exhibiting early sero-reversion. The random selection process of serum samples resulted in a pool of primary latitudinal sera (*i.e*., cross-sectional sera), while longitudinal sera (ideally from clinical trials of vaccine development) is required to catch the sero-conversion and sero-reversion phases. Furthermore, we intend to obtain a significantly larger number of samples to further study the aforementioned small probability events (Table S6). Finally, a field study at a large scale is needed to examine whether the probe combination of PPHM_covid-19_ calculated from a relatively small population is valid. Notably, our demonstration that sensitivity and specificity is population depended for whole-protein-based assays but not PPHMcovid-19 can be further confirmed with such work.

In conclusion, we completed the training and validation studies of PPHMcovid-19 for diagnosis of COVID-19. Given that PPHM has fixed cut-off values and that selection of apparently universal probes can be achieved with a small initial population, it is clear that PPHM development can be standardized, making it especially suitable for rapidly responding to sudden outbreaks of infectious diseases (*26, 27*). Moreover, the SANE protocol we demonstrate has the potential to significantly reduce the time required for quarantine for people with close contact history (*i.e*., from 14 days to 6 days). This PPHM platform is also able to provide a means of differentiating between infected and vaccinated hosts, which will be particularly useful during the development of a SARS-CoV-2 vaccine in the coming year.

## Data Availability

All data associated with this study are in the main text or Supplementary Materials.

## Materials and Methods

### Study design

A total of 973 serum samples were collected in this study from Chinese Center for Disease Control and Prevention and six hospitals, The Central Hospital of Wuhan (WCH), The First Affiliated Hospital of Guangzhou Medical University (FAH), The 2nd People’s Hospital of Shenzhen (SPH), Shenzhen Children’s Hospital (SCH), Jinling Hospital (JL) and Huoshenshan Hospital (HSS) respectively. Among them, 463 sera from 384 patients with COVID-19 symptoms (Table S1), and 55 sera from 48 patients without COVID-19 symptoms. 122 sera of hospital inpatients due to other diseases and 333 anonymous serum collected before 2019, of which 133 serum samples were from patients with influenza or systemic lupus erythematosus.

A total 518 serum samples of 432 patients were enrolled from two designated hospitals, WCH (415) and FAH (17). Among them, 80 patients from WCH had sequential serum samples. The median age of 432 enrolled patients was 60 years (IQR, 47 - 69 years) and 54.8% were females. Among them, 306 serum samples from 246 patients positive for SARS-CoV-2 with PCR on nasal and pharyngeal swab specimens, and another 159 serum samples from 139 patients were confirmed to be infected with SARS-CoV-2 by serological test according to the Guidelines of the Diagnosis and Treatment of New Coronavirus Pneumonia published by the National Health Commission of China (*6*).

Another 53 serum samples from 47 patients suspected to have COVID-19 but negative for SARS-CoV-2 with PCR for at least two sequential tests. Among them, 26 patients with suspected COVI-19 admitted to WCH with COVID-19 symptoms, and 3 close contacts (patients’ family member) without COVID-19 symptoms but abnormal pulmonary imaging.

The study was approved by the ethics committee of the four hospitals mentioned above (Medical Research Ethics No. 44, 2020). Written informed consent was waived in light of this emerging infectious disease of high clinical relevance. All healthy control subjects signed written informed consent prior to the collection of peripheral blood.

### Serological assays for SARS-CoV-2

All serum samples were inactivated at 56 °C for 30min and stored at −20 °C before testing. There are two serological assays in this study, single probe chemiluminescence assay (CLIA) and multi-probe PPHM assay.

Single probe CLIA was conducted on an Axceed 260 automatic CLIA analyzer (Bioscience (Tianjin) Co., Ltd.). The cut-off values of IgM and IgG are 53292 and 73400, respectively. The relative luminescence value (RLV) greater than or equal to 1.0 is positive for specific IgM and IgG.

Peptide-protein hybrid microarray (PPHM) included eight peptides and one whole protein (RBD) (GenScript, Jiangsu, China). Multi-probe PPHMCOVID-19 assay was conducted as previously described. Briefly, serum was first diluted 1:100 with serum-dilution buffer, then 100 μL diluted sample was added into each microarray well and incubated for 30 min or 2 hours on a shaker (500 rpm, 37°C). Then, the microarray incubated with horseradish peroxidase conjugated goat anti-human IgG (ZSGB-BIO, Beijing, China) for another 30 min on a shaker (500 rpm, 37°C). Finally, 1-step Ultra TMB-Blotting Solution (Thermo Scientific, USA) was used to detect the informative signal of IgGs against probes using microarray imager, which was then analyzed using the imager accompanied commercial software (Suzhou Epitope, Suzhou, China). The cut-off value of each probe is 10. Any two or more probes in PPHMcovid-19 whose signals are higher than 10 are confirmed as positive samples.

### Statistical analysis

The Fisher’s exact test was performed to identify statistically significant probes between negative and COVID-19 sera with a p-value cutoff of 0.05. Then the PPHMcovid-19 which contained eight peptides and one protein-RBD were calculated for combined diagnostic performance by an in-house script. The 95% CI of the sensitivity and specificity were calculated from binomial probabilities using adjusted Wald method. ROC was performed using R package “pROC”.

## One Sentence Summary

Guided by atypical humoral responses in subsets of COVID-19 patients that we detected based on an innovative serological testing technology, we have developed a diagnostic protocol that offers unprecedented performance for accurate diagnosis of COVID-19 in the clinic.

## Acknowledgments

This study was supported by Zhejiang University special scientific research fund for COVID-19 prevention and control (2020XGZX001 to N.Z., 2020XGZX025 to B.S., 2020XGZX040 to H.M.), China Evergrande Group (2020GIRHHMS04 to B.S. and H.M.) and Self-funded project of SINANO, CAS (E051020101 to H.M.).

## Author contributions

H.M., N.Z., B.S., and H.W. conceived the study; H.C., P.Z., D.G., S.H., D.M., R.W., J.H., Z.L., S.W., C.L. and X.X. performed all the experiments; H.C., Y.L., Y.D., and L.Y. analyzed the results; H.C., Y.L., W.X. and H.M. wrote the manuscript. All authors contributed to the revision and review of the manuscript.

## Data and materials availability

All data associated with this study are in the main text or Supplementary Materials.

## Supplementary Materials for

**DMI significantly reduces the sensitivity-and specificity-related impacts of variations in the particular population used for assay development**. In order to demonstrate the merit of using uniform cut-off value for individual probes, we listed response rates of all nine probes used in PPHMCOVID-19, as well as the N protein. Since the microarray can be used in a full PPHM configuration or as discrete ELISA probes, we used RBD and N protein in a PPHM as discrete ELISA probes. In general, the density curve for the response rate of negative sera for one probe has two peaks and a long tail toward the right direction (Fig. S1). The first and high peak reflects the fact that most sera showed a near zero signal, and the second and low peak indicates the level of nonspecific interaction (NSI). The central value of the second peak varies for different probes, and may be correlated to their “immunogenicity”, a general term used to describe/explain the ability/potential of a peptide/protein to activate the immune system/induce antibody production. The long tail toward the right side implies that NSI cannot be discriminated by signal (*17*).

**Fig. S1.**
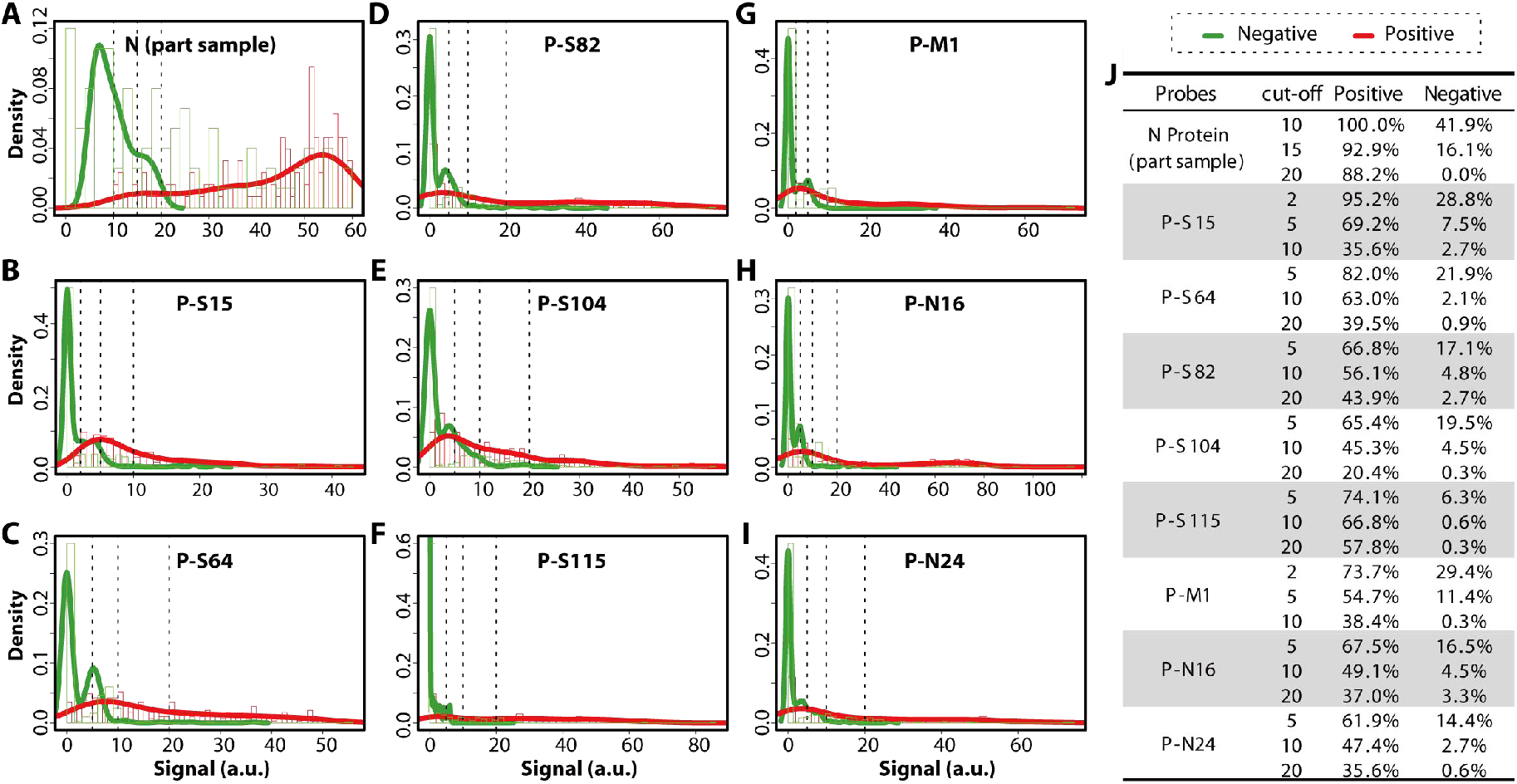
Impact of cut-off value on the sensitivity and specificity of whole protein-based and PPHM assays. (**A**) Anti-(N protein) IgG signals for a subgroup comprising 94 control and 128 COVID-19 sera. (**B**)-(**I**) Results of antibody testing of other probes for a training group of 333 control and 289 COVID-19 serum samples. (**J**) List of response rates for different probes under different cut-off values.

One of the reasons that cause sampling problems maybe the variety of conditions triggering hospital visit in daily practice, *i.e*., the various combinations of viral-status, sero-status, and disease-status upon sampling for clinical tests. For example, at sampling-4, Hosts 3 and 4 will have positive test results; Hosts 1 and 2 will have false negatives due to sampling in their window period; and Host 5 will have negative test results because sampling is conducted in the sero-negative period. However, at sampling-9, although most cohort samples will test positive due to overlap in the prolonged sero-positive period, some hosts, like Host 2, will show positive results due to sero-conversion, while others, *e.g*., Host 4, will have negative test results due to sero-reversion.

There are three goals to achieve, determining i) the presence of SARS-CoV-2; ii) the presence of anti-(SARS-CoV-2) IgG; iii) COVID-19. PCR-based assays can determine whether there is active SARS-CoV-2 infection. Since active SARS-CoV-2 infection is a necessary and sufficient condition for being COVID-19 patient, positive PCR test results can be used to diagnose COVID-19. However, a host could be a COVID-19 patient after SARS-CoV-2 clearance, *i.e*., negative PCR test result due to sampling in viral-absence period would deliver a false negative CVOID-19 result.

Similarly, negative serological assay (PPHMCOVID-19 or CLIA) results due to sampling in the IgG production window period of SARS-CoV-2 infection are true negative in term of IgG production, however, are false negative results for COVID-19 diagnosis. **Thus, we define all test results referring to COVID-19 diagnosis unless otherwise indicated**.

**Fig. S2.**
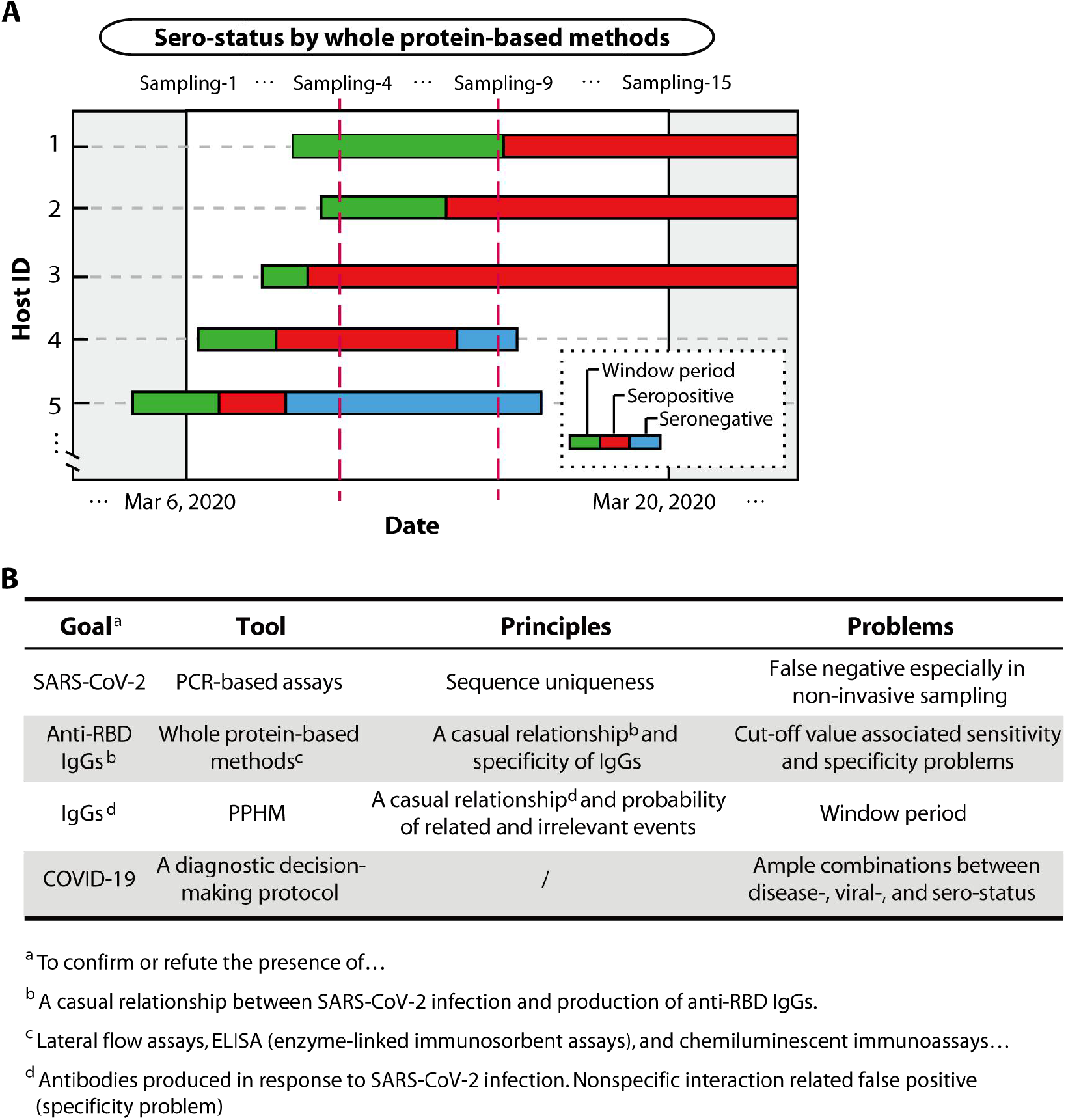
Diagnostic tools need to be used under proper conditions. (**A**) Schematic illustration of serum sampling marked by date. Cohort sera collected at any given date will contain samples from all three stages of sero-status, regardless of whether the dpo is identical among samples. (**B**) Different diagnostic goals should be selected for different detection tools.

Of the 414 confirmed COVID-19 patients, we had access to sequential serum samples for 80. Among these, 71 patients were sampled within their sero-positive period.

**Fig. S3.**
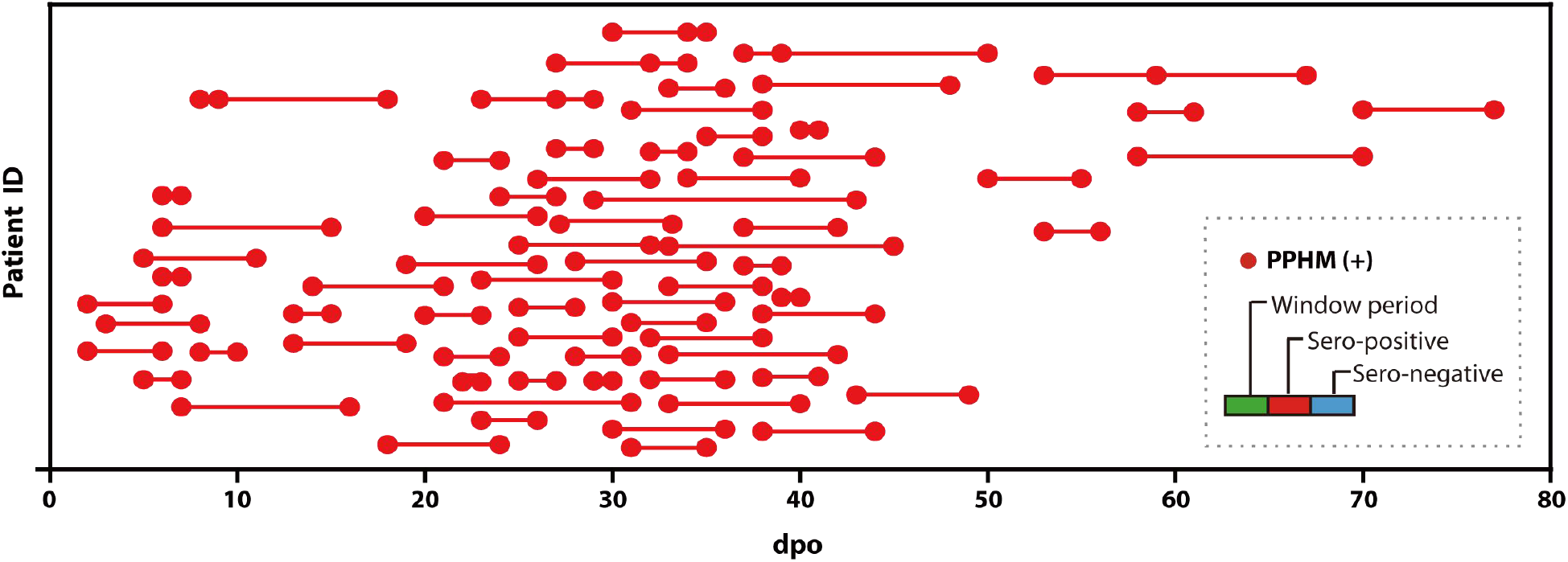
Among the 80 patients with sequential serum samples available, 71 patients were sampled in their sero-positive period.

We found seven serum samples (dpo > 10) that have clear PCR negative time. Among them, three serum samples (Serum-4-11dpo, Serum-85-27dpo, and Serum-132-42dpo) were collected on the day or after the day that PCR turned into negative, which we suspected are the sera from patients characterized by anti-RBD IgG fast disappearing. Serum-41-15dpo and Serum-64-14dpo, which showed PPHMCOVID-19 positive and RBD negative results while PCR still positive, may be caused by PCR testing not frequent enough so that the exact date of viral clearance should be earlier than the serum sampling time. In contrast, Serum-48-40dpo and Serum-132-37dpo with the late dpo, are easily determined to be in their sero-negative period, and PCR testing were not frequent enough. Therefore, we proposed these seven serum samples were not in the window period of RBD, and such results indicated pathogen-derived peptides have longer sero-positive period than RBD (whole protein).

**Fig. S4.**
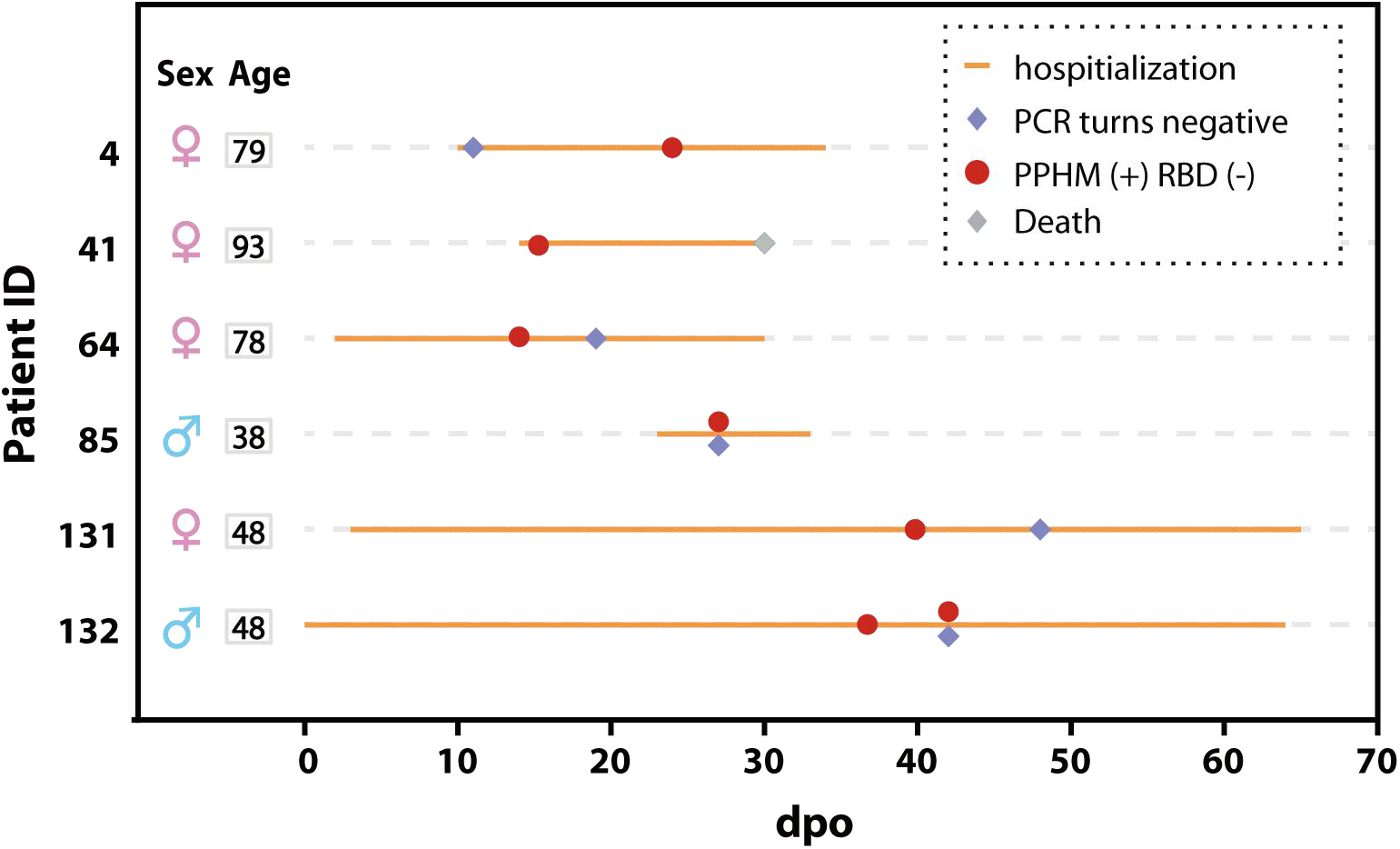
Basic information of the 7 serum samples (dpo > 10) from 6 COVID-19 patients exhibited positive PPHM_COVID-19_ results but negative RBD signal.

The rationale underlying the design of the high sensitivity iteration of PPHM_COVID-9_ assay, was based on two accepted principles of immunology. i) IgGs produced in the window period are low in both concentration and affinity; ii) IgGs produced around the sero-reversion point are also low in concentration but exhibit high affinity. Based on the observation that IgGs with high affinity but low concentration (cognate Ab-Ag pairs) can be detected after a prolonged chip incubation with sera, while the detection of low affinity IgGs (non-specific interaction Ab-Ag pairs) is not affected by incubation time, we designed the high-sensitivity iteration of PPHMCOVID-9 test to diagnose serum samples collected either in the window period or around the sero-reversion point. For example, Patient-165, 42, and Neg1 that were in sero-negative period, window period, and confirmed other etiology, respectively, were tested using the high-sensitivity iteration of PPHM_COVID-9_. Patient-165 at 27 dpo showed an enhanced response of two probes (P-S15 and PS64), *i.e*., DMI changed from 0 to 2. Patient-Neg1 with other etiology remained as negative under both conditions, supporting our design rationale. Further, these results offer experimental confirmation that longer incubation durations can indeed increase the sensitivity of PPHM probes.

**Fig. S5.**
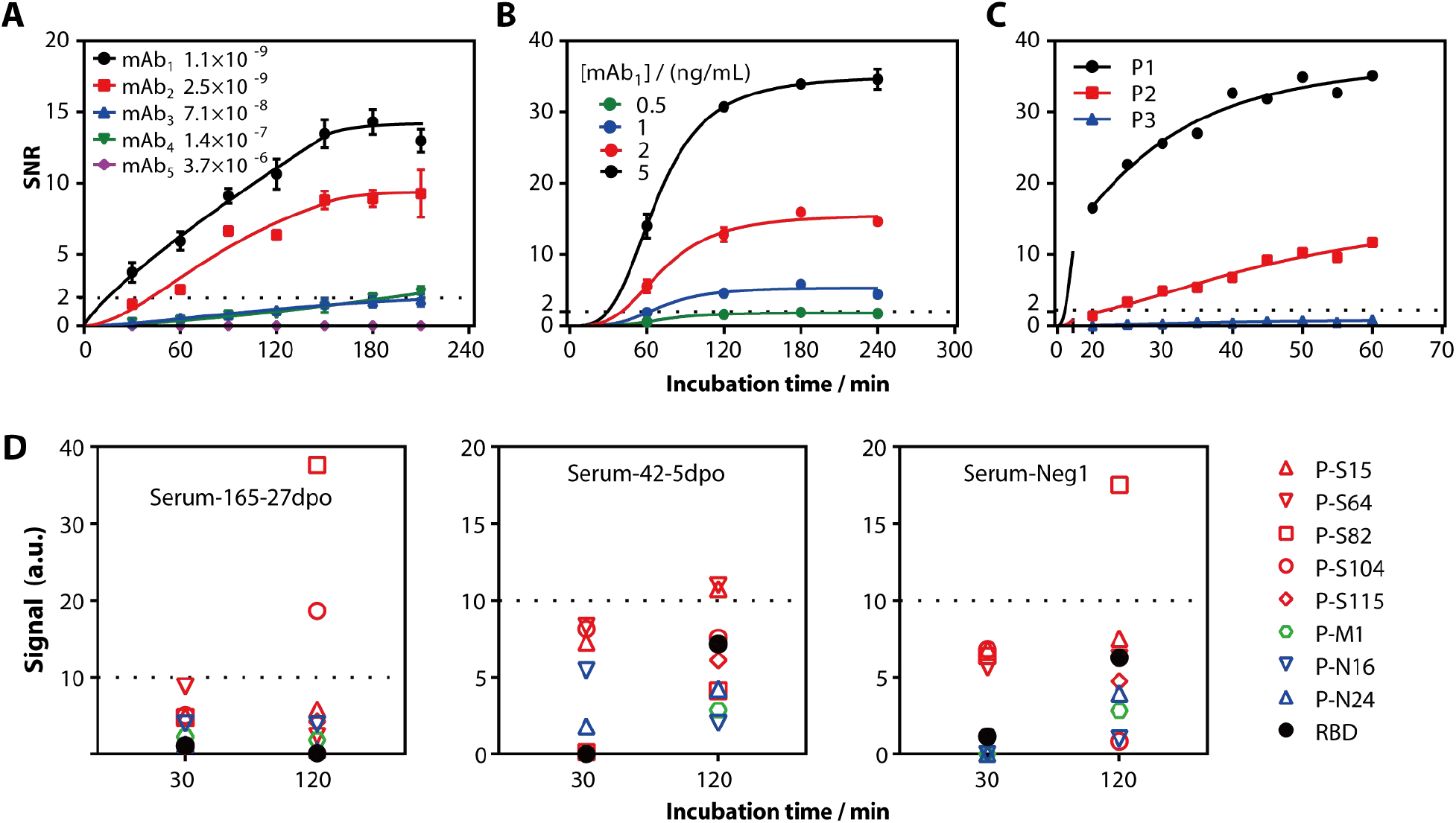
Prolonging the incubation duration to improve the performance of PPHM detection. We obtained five monoclonal antibodies (mAb1~mAb5) with different affinity for the same short peptide P1 (DQPQNLEEILMHCQT) by the classical mouse hybridoma method. (**A**) At the same antibody concentration (5 ng/mL), the response characteristics of different affinity mAbs are different. (**B**) At different concentrations of the same mAb (*i.e*., the same affinity), the response characteristics are different. (**C**) Demonstrates the response of mAb1 to cognate short peptide P1 and non-cognate short peptides P2 and P3. (**D**) Example: Patient-165 in late dpo has three probes converted from negative to positive, Patient-42 in early dpo has two probes converted, and Patient-Neg1 of other etiology is negative for all probes. After prolonging the incubation time One probe turns positive, but does not affect the detection result (still negative). The dotted line indicates the cut-off value.

To investigate whether PPHM_COVID-9_ testing could help identify patients with COVID-19, we screened 29 suspected subjects, with 26 who displayed symptoms of COVID-19 and another 3 without symptoms of COVID-19 yet who showed abnormal radiological findings; all of these subjects were viral RNA negative based on 111 testing; some showed more than one negative test. Notably, all 29 subjects showed positive PPHM_COVID-19_ results.

**Fig. S6.**
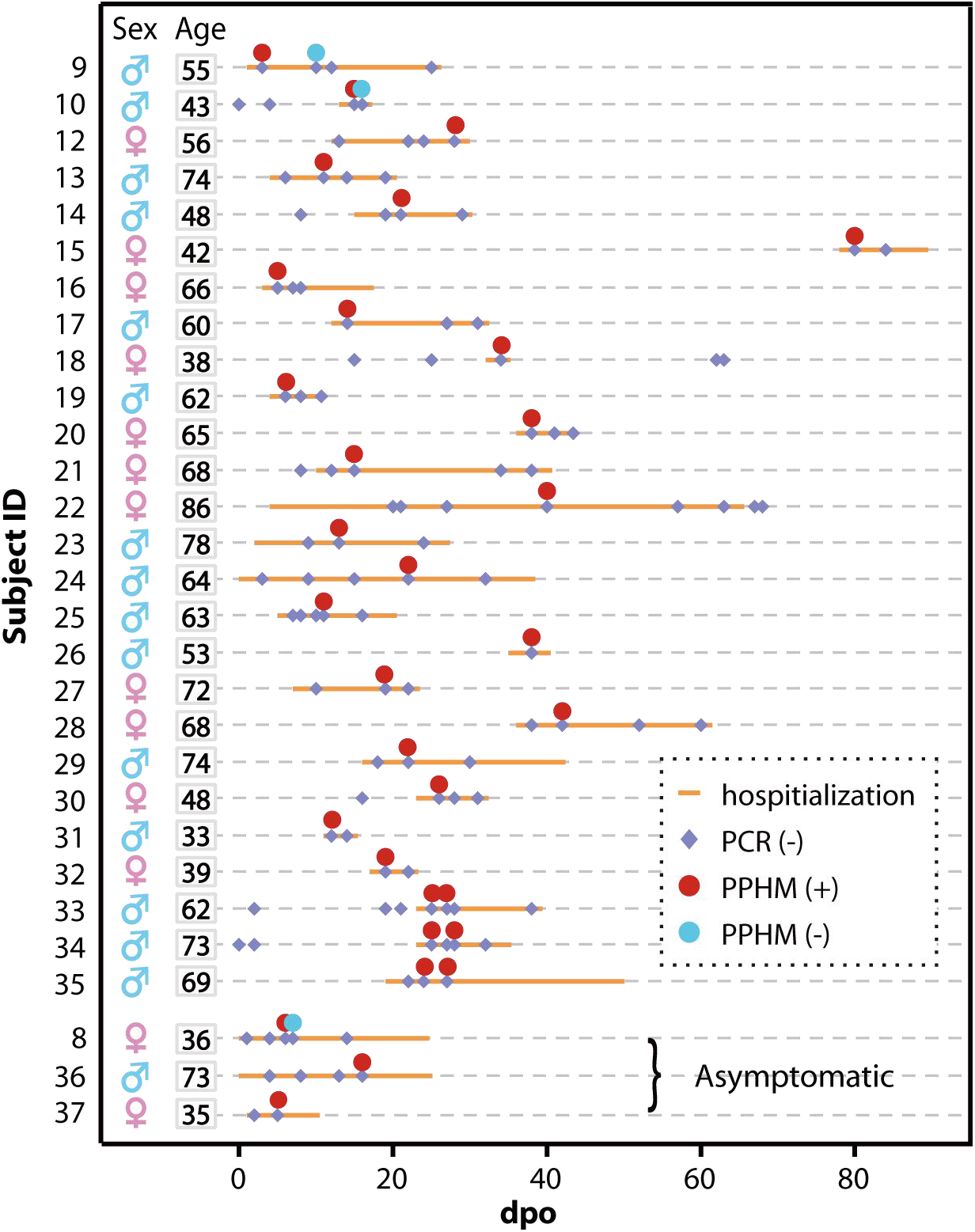
Basic information of the 35 suspected serum samples from 29 suspected subjects.

**Fig. S7.**
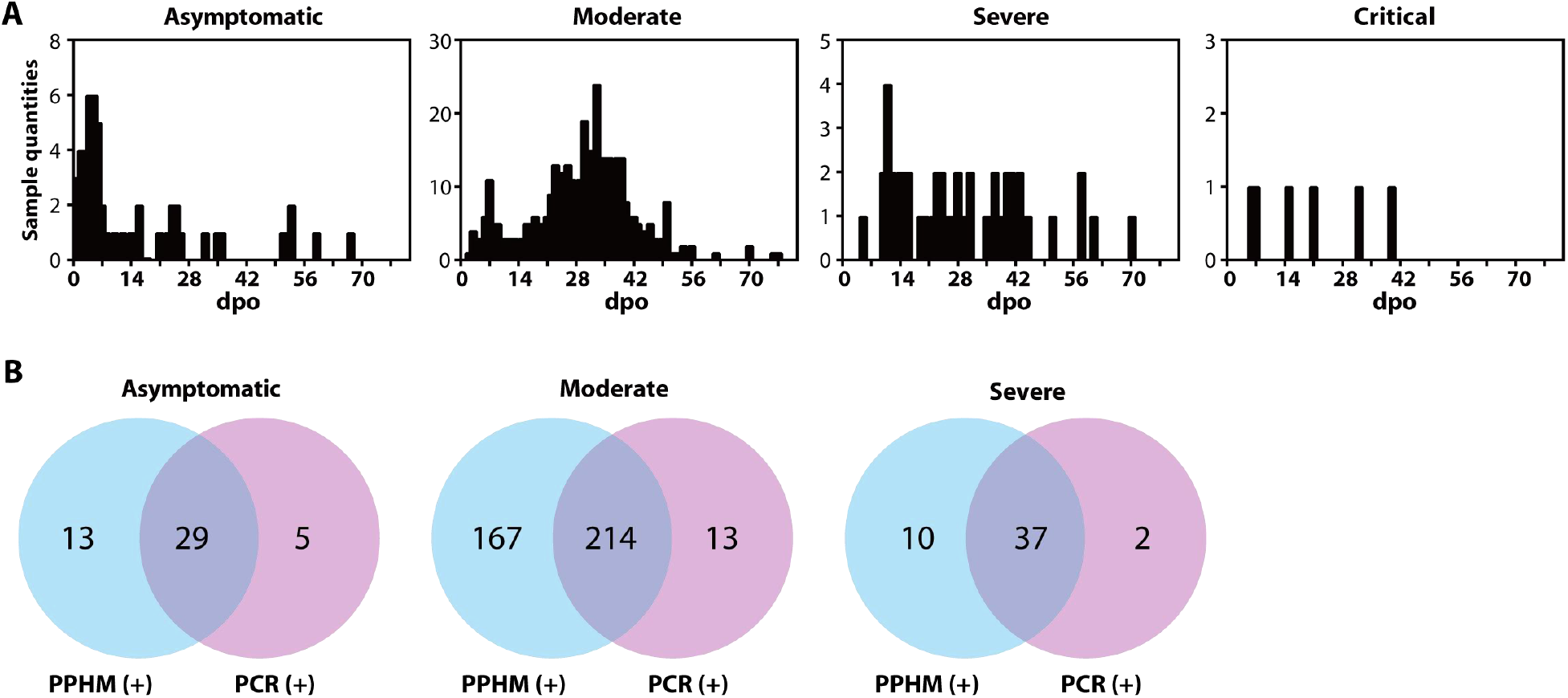
For asymptomatic, moderate and severe patients, the detection performance of PPHM_COVID-19_ is superior compared to a whole-protein (RBD)-based assay and a PCR assay. (**A**) The dpo distribution of four clinical patients. (**B**) Comparison of PPHM and PCR detection results as Venn diagrams for asymptomatic, moderate, and severe patients.

There are three scenarios in this study that require different dpo determination. The diagnosis is initiated i) by the subject him/herself upon the onset of symptoms. When the subject is confirmed as COVID-19 by PCR-based assay or other criterion, the dpo is recorded based on self-reporting from the patient (Fig. S8, black line). ii)-iii), by the subject upon confirmation with close contact history. According the guideline V7, if the subject is confirmed by positive PCR result, the date first positive PCR test result obtained is determined as 1 dpo, thus dpo is relative uncertain (Fig. S8, blue line). If a negative PCR test result but positive radiology result, the date first positive radiology result obtained is determined as 1dpo (Fig. S8, green line). If the dpi date could be affirmatively confirmed, dpo of three scenarios could be compared (Fig. S8, red line).

**Fig. S8.**
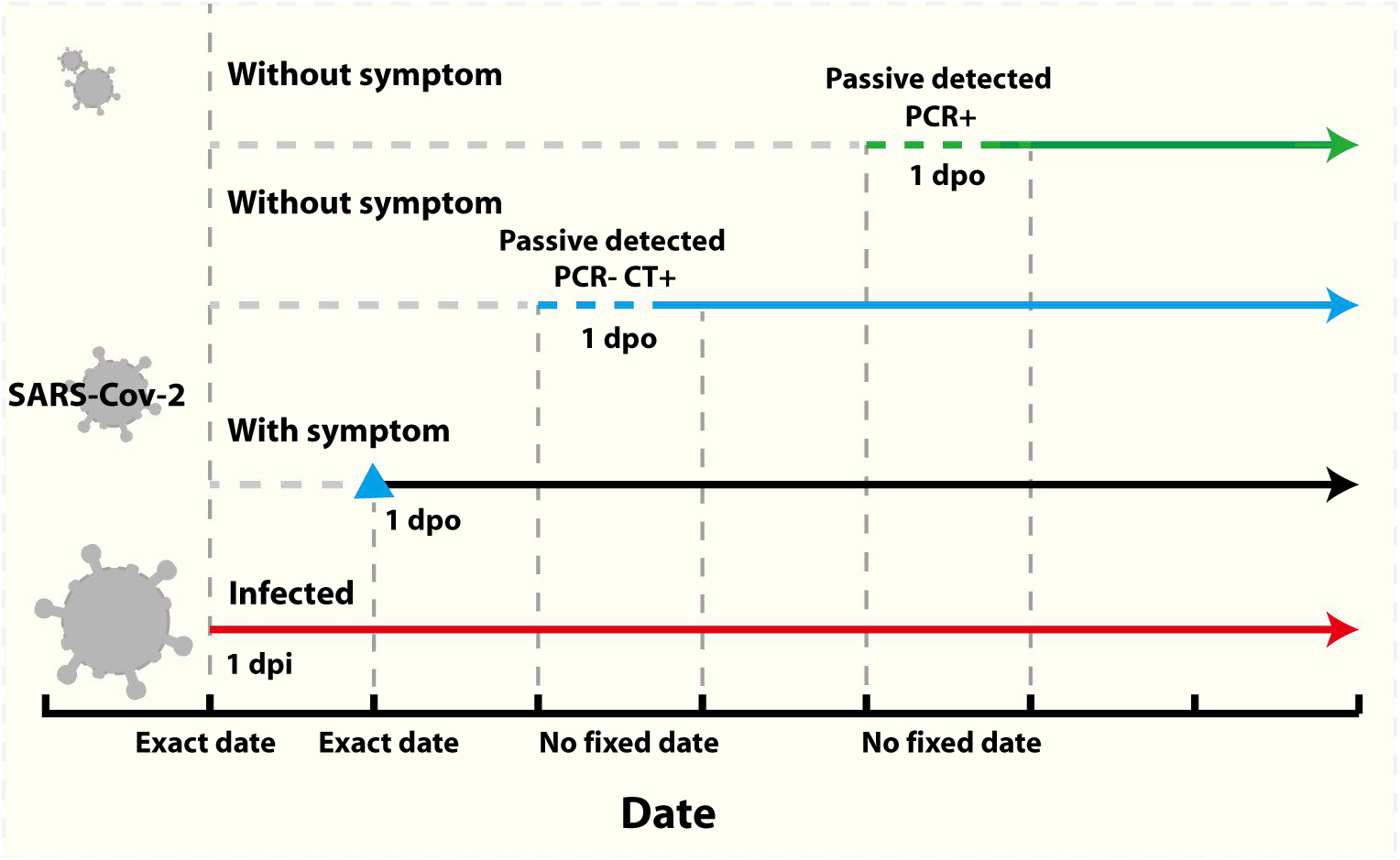
Three different dpo determination scenarios for symptomatic and asymptomatic patients.

**Fig. S9.**
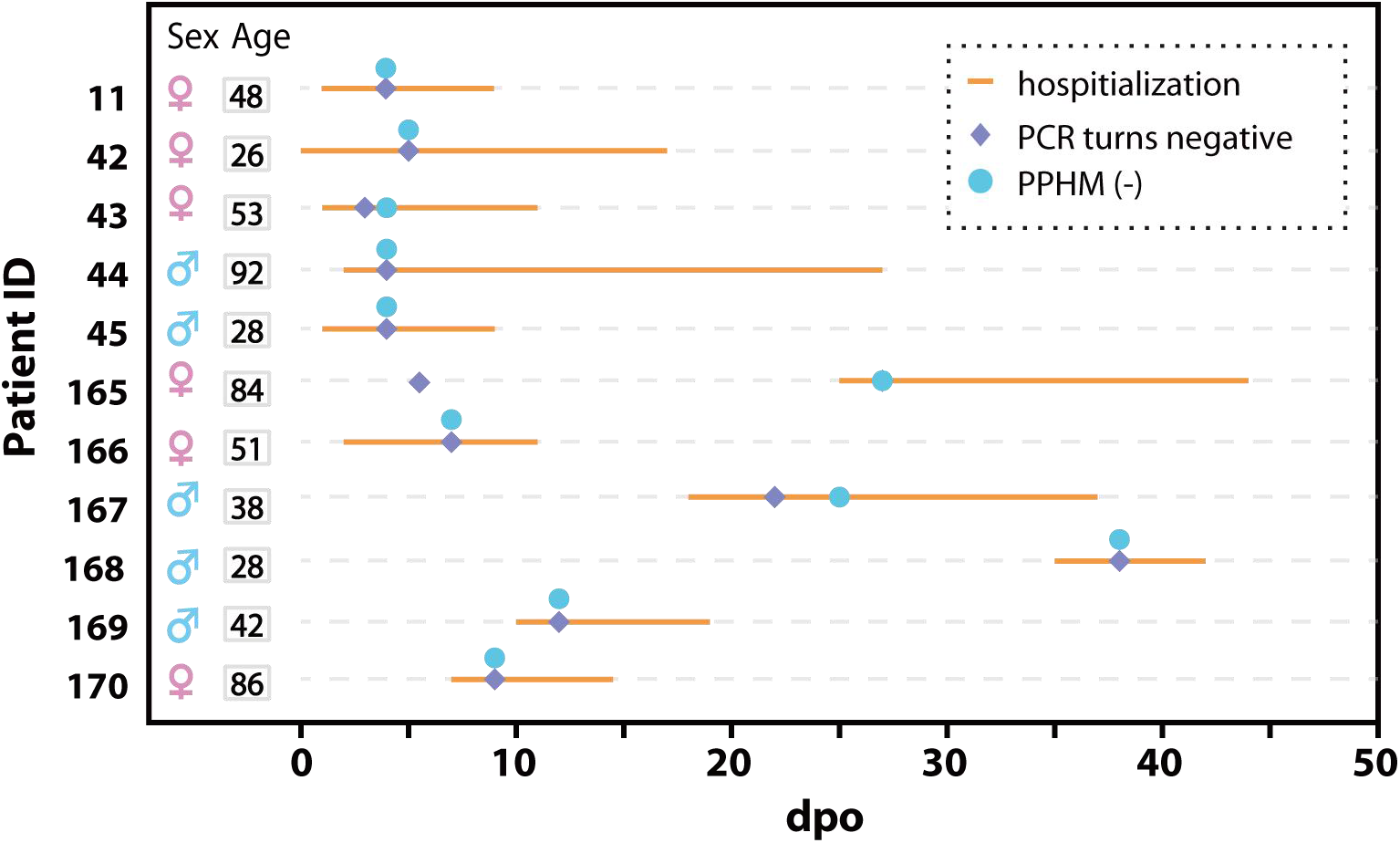
The PPHM_COVID-19_ diagnostic results for a patient subgroup of early sero-reversion.

**Fig. S10.**
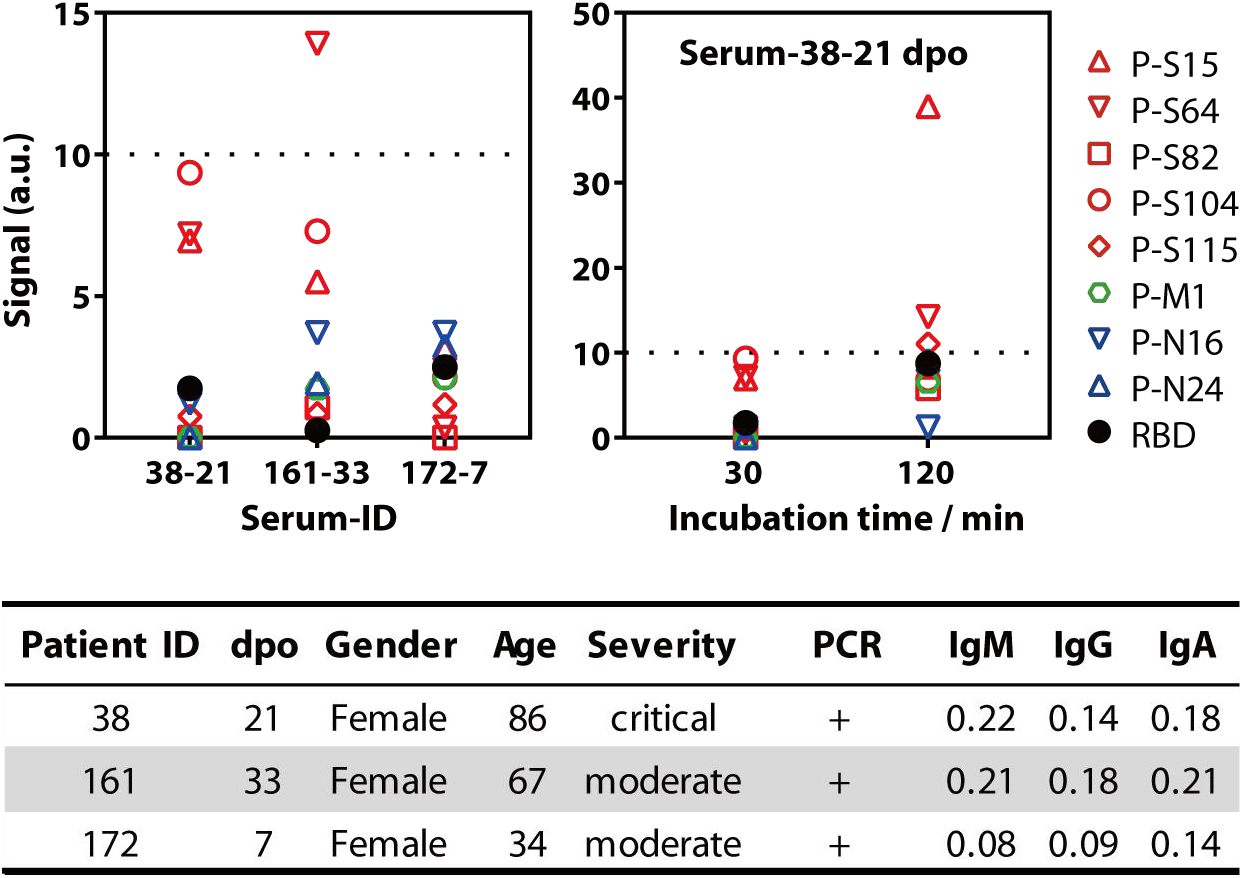
Basic information of three patients exhibited negative PPHM_COVID-19_ but with paired positive PCR test results.

From two patients with sequential serum samples, we observed that anti-peptide IgGs entered into the sero-positive period while RBD is still in its window period.

**Fig. S11.**
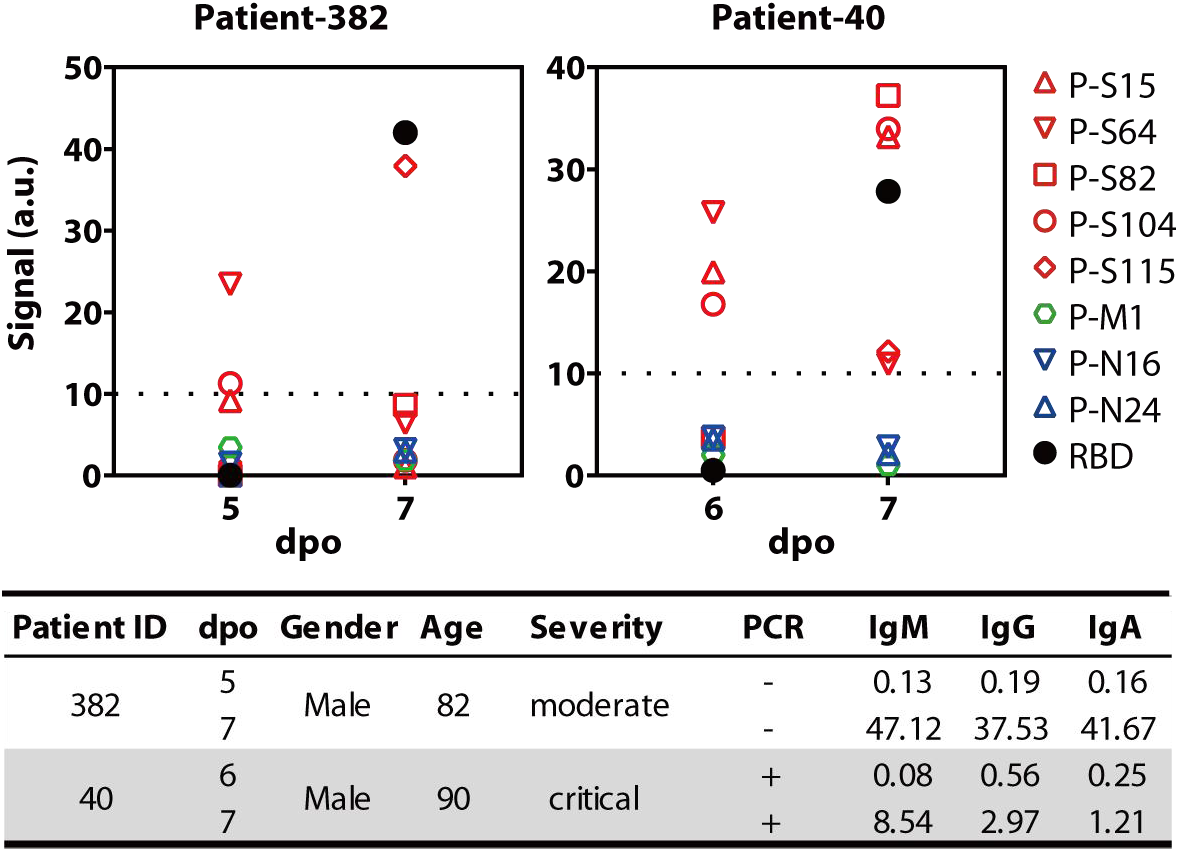
Some peptides enter sero-positive period while RBD is still in its window period.

**Fig. S12.**
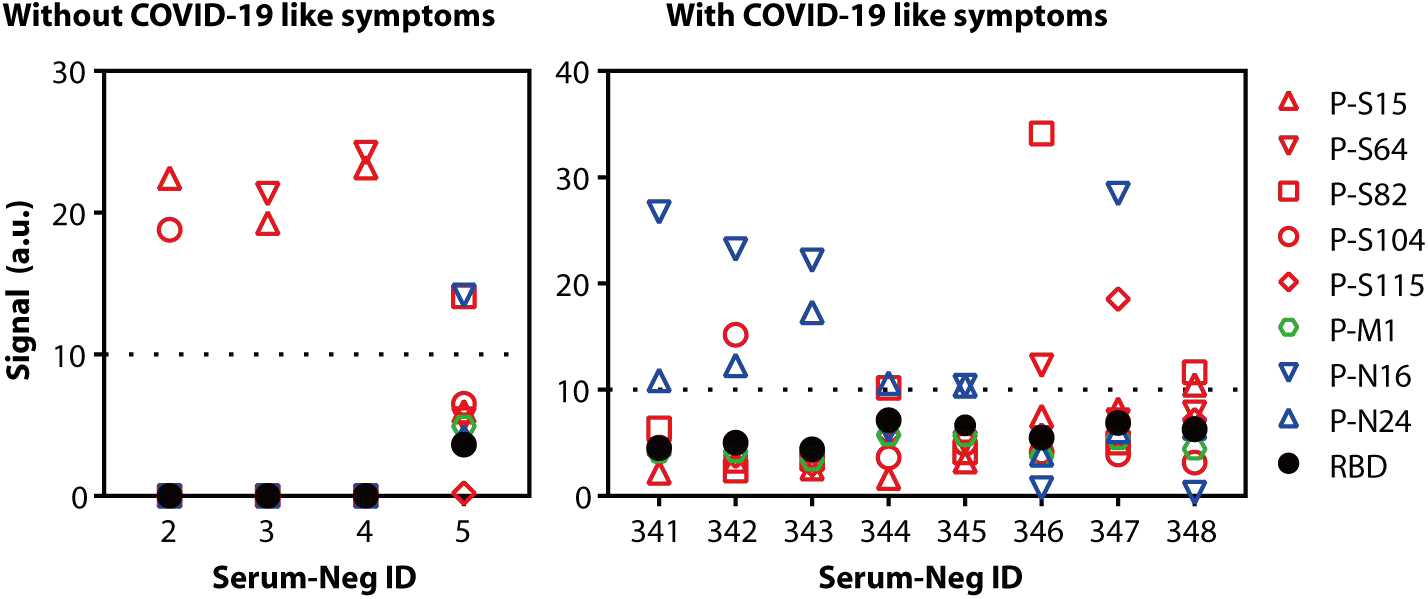
PPHM_COVID-19_ diagnostic results of 12 false positive serum samples. All showed negative anti-RBD IgG results.

Of the 473 total control serum samples, 12(12/473, 2.5%) showed false positive PPHM results (Fig. S12). All twelve serum samples can be determined as false positive through RBD negative in addition to epidemiological history. Of the nine, six were SLE patients, three were healthy, in agreement with our previous results supporting that it is not possible to predict when and where NSI/USI will happened, thus posing serious problems to present serological assays (*17*). PPHM can reduce the frequency by using multiple probes. N protein is less specific thus is not used. As SARS-CoV-2 continues to spread, the specificity of RBD is expected to decrease, implying RBD may not be suitable for future diagnosis of COVID-19.

We demonstrated, through the comparison of two SANE protocols that incorporated PPHM_COVID-19_ and CLIA, respectively, that PPHM_COVID-19_ is essential for achieving full sensitivity and specificity. For outpatients with COVID-19-like symptoms, while CLIA is incapable of distinguish false positive and positive testing results resulting in 13 ambiguity cases (Fig. S13B), PPHM_COVID-19_ can discriminate false positive from positive resting results during the continuous monitoring step because false positive will remain RBD negative (Fig. S13A). Although in SANE protocol using PPHM_COVID-19_ will have more enrollments than using CLIA, i.e., 57 vs 40, these outpatients will need self-isolation or quarantine anyway. The reason we can determine COVID-19 based on PPHM_COVID-19_ (DMI > 2) and RBD double positive is that RBD now has extremely high specificity so for one negative serum to be both PPHM_COVID-19_ and RBD false positive is almost impossible (3 independent events to simultaneously happen). Similarly, for outpatients without COVID-19-like symptoms, SANE protocol using CLIA produced 2 ambiguity cases (Fig. S13D) that are both confirmed as COVID-19 cases by the SANE protocol using PPHMCOVID-19 (Fig. S13C).

**Fig. S13.**
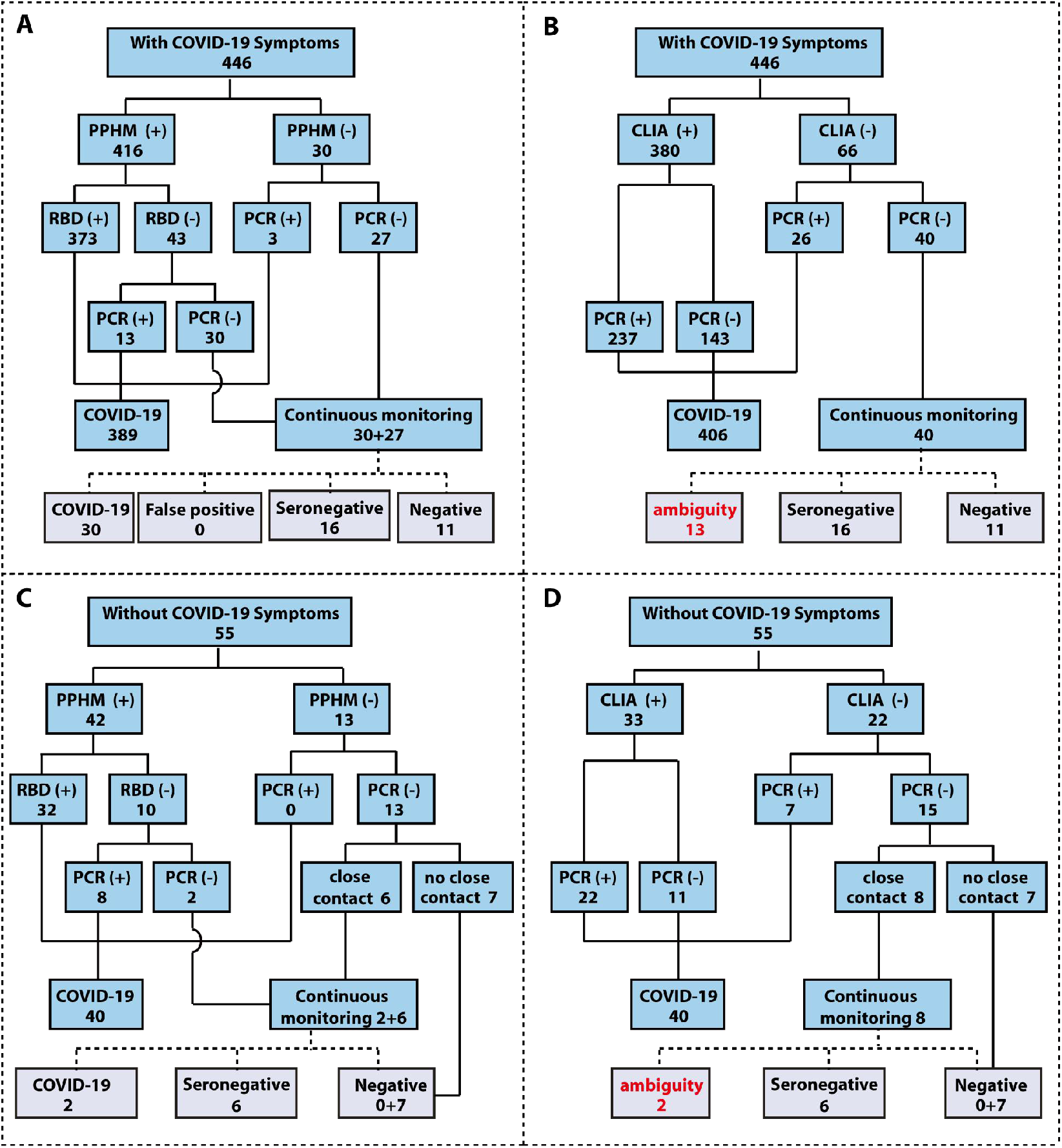
Comparison of performance of SANE protocols that incorporates either PPHM_COVID-9_ or CLIA. (**A**) The result of SANE protocol combined PPHM_COVID-19_ and PCR in 446 serum samples from the patient with COVID-19 like symptoms. (**B**) Matched CLIA results. (**C**) The result of SANE protocol combined PPHM_COVID-19_ and PCR in 55 serum samples from the patient without COVID-19 like symptoms. (**D**) Matched CLIA results.

**Table S1.**
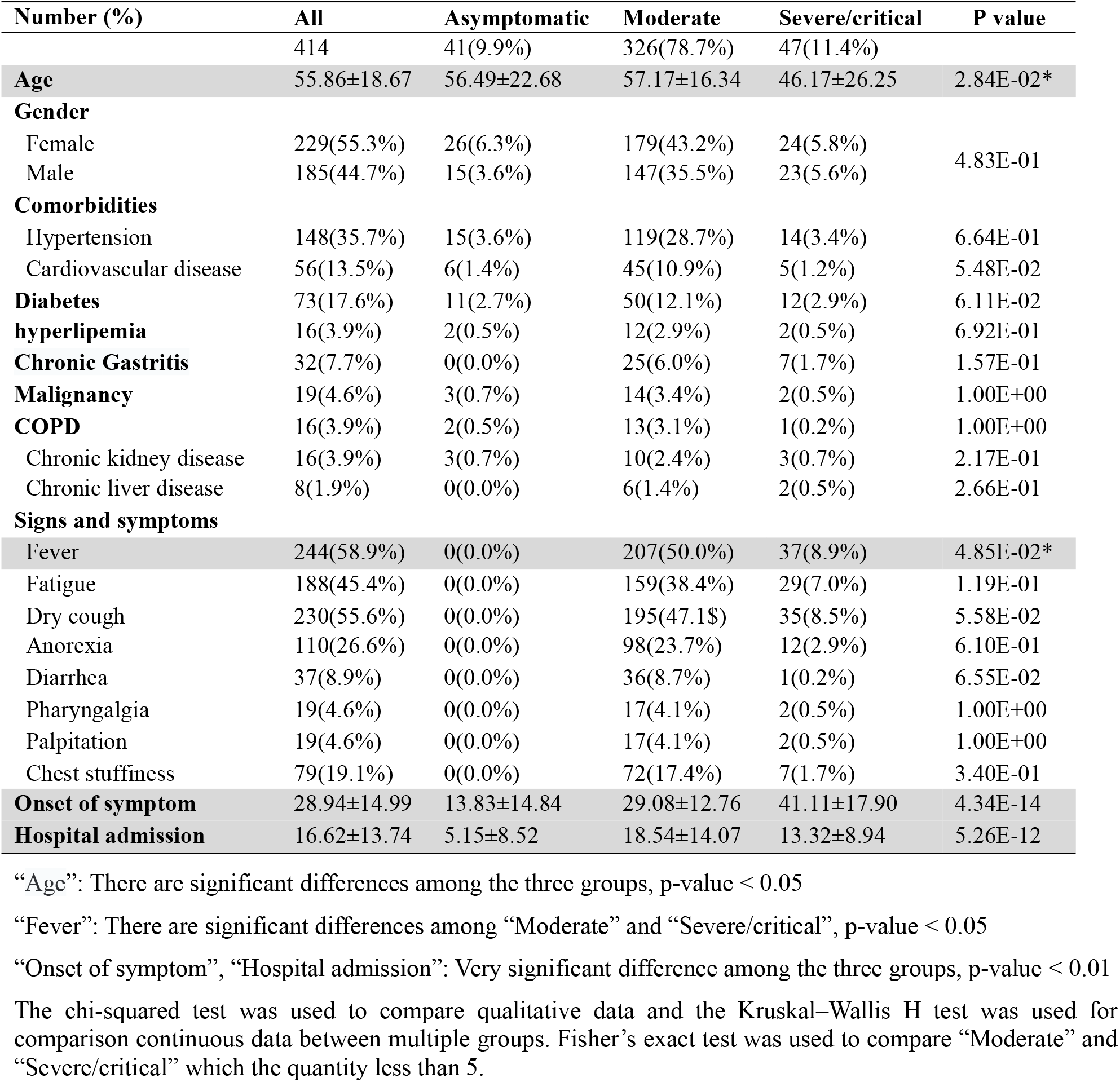
Baseline characteristics of patients from the cross-sectional study.

**Table S2.** Amino acid sequence of eight probes for PPHM_COVID-19_.

| Probe | Amino acid sequence | Region |
| --- | --- | --- |
| P-S15 | LGVYYHKNNKSWMESEFRVY | 141-160 |
| P-S64 | PTWRVYSTGSNVFQTRAGCL | 631-650 |
| P-S82 | KPSKRSFIEDLLFNKVTLAD | 811-830 |
| P-S104 | ECVLGQSKRVDFCGKGYHLM | 1031-1050 |
| P-S115 | LQPELDSFKEELDKYFKNHT | 1141-1160 |
| P-M1 | MADSNGTITVEELKKLLEQW | 1-20 |
| P-N16 | PANNAAIVLQLPQGTTLPKG | 151-170 |
| P-N24 | ESKMSGKGQQQQGQTVTKKS | 231-250 |

PPHM_COVID-19_ has eight peptide probes and one protein-RBD probe, which exhibited negative response rates ranging from 0.3% to 5%, and positive response rates ranging from 36% to 85% (Fig. 1H). For the 333 control serum samples, P-S82, P-S104, and P-N16 have 16(5%), 15(5%), and 15(5%) responding samples, respectively (Fig. 1H).

For DMI = 2, there are 36 possible combinations (Table S3). The predicted values of three combinations, namely (P-S82, P-S104), (P-S82, P-N16), and (P-N16, P-S104), are all roughly 0.25% (1/333). The experimental values are all roughly 0.9% (3 or 2/333). The overall specificity and sensitivity for DMI = 2 are 2.7% (9/333), which is lower than negative response rates of five probes, and 92% (267/289), which is higher than positive response rates of all nine response rate.

**Table S3.** All 36 possible two-probe combinations of nine probes in our PPHM_COVID-19_.

| DMI=2 | Control (n=333) |  | Positive (n=289) |  |
| --- | --- | --- | --- | --- |
| Response | Number | Rate | Number | Rate |
| P-S15, P-S64 | 2 | 1% | 92 | 32% |
| P-S15, P-S82 | 0 | 0% | 57 | 20% |
| P-S15, P-S104 | 1 | 0% | 92 | 32% |
| P-S15, P-S115 | 0 | 0% | 68 | 24% |
| P-S15, P-M1 | 0 | 0% | 37 | 13% |
| P-S15, P-N16 | 0 | 0% | 55 | 19% |
| P-S15, P-N24 | 0 | 0% | 50 | 17% |
| P-S15, RBD | 0 | 0% | 84 | 29% |
| P-S64, P-S82 | 0 | 0% | 106 | 37% |
| P-S64, P-S104 | 0 | 0% | 112 | 39% |
| P-S64, P-S115 | 0 | 0% | 127 | 44% |
| P-S64, P-M1 | 0 | 0% | 75 | 26% |
| P-S64, P-N16 | 0 | 0% | 98 | 34% |
| P-S64, P-N24 | 0 | 0% | 96 | 33% |
| P-S64, RBD | 0 | 0% | 161 | 56% |
| P-S82, P-S104 | 0 | 0% | 68 | 24% |
| P-S82, P-S115 | 0 | 0% | 132 | 46% |
| P-S82, P-M1 | 0 | 0% | 85 | 29% |
| P-S82, P-N16 | 1 | 0% | 96 | 33% |
| P-S82, P-N24 | 1 | 0% | 97 | 34% |
| P-S82, RBD | 0 | 0% | 161 | 56% |
| P-S104, P-S115 | 0 | 0% | 92 | 32% |
| P-S104, P-M1 | 0 | 0% | 50 | 17% |
| P-S104, P-N16 | 1 | 0% | 62 | 21% |
| P-S104, P-N24 | 1 | 0% | 62 | 21% |
| P-S104, RBD | 0 | 0% | 113 | 39% |
| P-S115, P-M1 | 0 | 0% | 101 | 35% |
| P-S115, P-N16 | 0 | 0% | 116 | 40% |
| P-S115, P-N24 | 0 | 0% | 116 | 40% |
| P-S115, RBD | 0 | 0% | 192 | 66% |
| P-M1, P-N16 | 0 | 0% | 71 | 25% |
| P-M1, P-N24 | 0 | 0% | 66 | 23% |
| P-M1, RBD | 0 | 0% | 111 | 38% |
| P-N16, P-N24 | 3 | 1% | 86 | 30% |
| P-N16, RBD | 0 | 0% | 133 | 46% |
| P-N24, RBD | 1 | 0% | 135 | 47% |

Three subgroups were randomly selected from the training group. Subgroup-1, −2, and −3 comprised (control, COVID-19) sera at (50, 50), (100, 100), and (150, 150), respectively.

**Table S4.** Impacts of variations in the population used for assay development on response rates for probes and PPHM. **(A)** List of response rates for PPHM for the three subgroups. (**B**) List of sensitivity and specificity values under different DMI for the three subgroups.

| Response rate | subgroup-1 |  | subgroup-2 |  | subgroup-3 |  |
| --- | --- | --- | --- | --- | --- | --- |
|  | Positive | Control | Positive | Control | Positive | Control |
| P-S15 | 30% | 2% | 32% | 3% | 33% | 1% |
| P-S64 | 64% | 4% | 59% | 2% | 67% | 2% |
| P-S82 | 66% | 6% | 51% | 4% | 58% | 4% |
| P-S104 | 50% | 8% | 46% | 4% | 42% | 2% |
| P-S115 | 72% | 0% | 69% | 1% | 66% | 1% |
| P-M1 | 42% | 0% | 41% | 0% | 39% | 0% |
| P-N16 | 54% | 8% | 48% | 3% | 49% | 5% |
| P-N24 | 56% | 6% | 38% | 5% | 52% | 3% |
| RBD | 92% | 0% | 82% | 2% | 83% | 0% |

**(B)** List of sensitivity and specificity values under different DMI for the three subgroups.
| cut-off | subgroup-1 |  | subgroup-2 |  | subgroup-3 |  |
| --- | --- | --- | --- | --- | --- | --- |
|  | Sensitivity | Specificity | Sensitivity | Specificity | Sensitivity | Specificity |
| DMI $\geq 1$ | 98% | 74% | 95% | 81% | 95% | 84% |
| DMI $\geq 2$ | 96% | 94% | 92% | 96% | 93% | 98% |
| DMI $\geq 3$ | 92% | 98% | 82% | 99% | 87% | 100% |
| DMI $\geq 4$ | 76% | 100% | 70% | 100% | 73% | 100% |
| DMI $\geq 5$ | 64% | 100% | 47% | 100% | 55% | 100% |
| DMI $\geq 6$ | 48% | 100% | 37% | 100% | 41% | 100% |
| DMI $\geq 7$ | 32% | 100% | 27% | 100% | 26% | 100% |
| DMI $\geq 8$ | 16% | 100% | 13% | 100% | 14% | 100% |
| DMI $\geq 9$ | 4% | 100% | 3% | 100% | 4% | 100% |

**Table S5.** Three serum samples were PPHM_COVID-19_ positive but RBD negative, and CLIA were all borderline positive.

| Serum ID | P-S15 | P-S64 | P-S82 | P-S104 | P-S115 | P-M1 | P-N16 | P-N24 | RBD | IgM | IgG | IgA |
| --- | --- | --- | --- | --- | --- | --- | --- | --- | --- | --- | --- | --- |
| 59-5 | 27.74 | 20.83 | 2.56 | 31.59 | 1.85 | 0.86 | 11 | 2.16 | 0 | 0.07 | 1.05 | 0.21 |
| 91-3 | 21.02 | 37.75 | 0.6 | 35.18 | 1.44 | 0.42 | 4.68 | 1.54 | 3.44 | 0.07 | 1.04 | 0.19 |
| 124-5 | 37.85 | 36.96 | 7.6 | 29.38 | 3.02 | 3.08 | 6.15 | 5.94 | 2.38 | 2.19 | 1.4 | 0.28 |

In order to ensure the integrity of the PPHM analyses and to further explore the causes of false negatives and false positives, we categorized 833 tested serum samples into 28 theoretically possible combinations that encompass the four possible combinations of PCR-based and PPHM test results that can result from the seven possible combinations of virus infection and antibody states. Of the 19 combinations for which no serum samples were identified in this cohort: Theoretical category ➄ patients (No. 17-20) represent those who have received immediate drug administration upon virus infection; eight groups (No. 1, 2, 5, 13, 21, 22, 25, 26) include all possible false positives; seven groups (No. 7, 8, 12, 15, 16, 24, 27) include all possible scenarios leading to false negative results. While it is the case that false positive PCR results have been reported during this pandemic (*9*), the low probability of these events is supported by the results observed in this cohort. The high prevalence of false negative results observed in PCR-based assays during the current pandemic, viewed alongside our finding of low prevalence for these combinations in this cohort (No. 7 and 15), together supports the low probability of PPHM false positive results. This robust avoidance against false positive results was also suggested by the absence of group No. 27 samples in this cohort. False negative results such as in groups No. 8, 12, 16, and 24; these are all theoretically possible, but were not observed, most likely due to the cohort size.

Of the eight combinations with serum samples identified in this study, Combination No. 9 in category ➂ accounted for the largest portion of positive serum samples; this combination contains the highest probability events. Combination No. 11 includes 178 serum samples with PCR negative but PPHM positive results, which illustrate the advantage of PPHM tests over PCR-based assays. This high prevalence of false negative PCR-based results is attributable to high sampling variation for this testing method, in contrast to serological sampling. Combination No. 6 contains 11 patients that were sampled during the window period of RBD antibody production, but were identified as seropositive by PPHM_COVID-19_, indicating that PPHM_COVID-19_ has improved sensitivity for samples collected at early dpo. Combination No. 28 includes eight patients that exhibited rapidly disappearing antibodies. Combination No. 4, in category ➀; accommodates the largest portion of negative serum samples (473 out of 973). Combination No. 3 includes only one PPHM false positive serum sample from Subject 1, which was corrected by simple, auxiliary confirmation tests: Subject 1 was positive for two short peptide probes but had no response to the RBD probe, and no close contact history, indicating a high probability of a false positive by PPHM-1 due to high [mAb].

**Table S6.** Twenty-eight combinations of virus presence, antibody production, PCR and PPHM test results.

| Real status<br>(virus, antibody) |  | No.<br>Group | Test results |  | Characteristic/Explanation | In our cohort |
| --- | --- | --- | --- | --- | --- | --- |
|  |  |  | PCR | PPHM |  |  |
| ① | (-, -)<br>never infected | 1 | + | + | PCR false positive; high [mAb] causes PPHM false positive | Not found |
|  |  | 2 | + | - | PCR false positive | Not found |
|  |  | 3 | - | + | High [mAb] causes PPHM false positive (other etiology) | SLE patients |
|  |  | 4 | - | - | Healthy people who have never been infected | Healthy people |
| ② | (+, -)<br>viral presence,<br>window period | 5 | + | + | Infected and high [mAb] causes PPHM false positive | Not found |
|  |  | 6 | + | - | IgGs stay in window period (mostly dpo ≤10) | 11 |
|  |  | 7 | - | + | PCR false negative due to non-invasive method and high [mAb] causes PPHM false positive | Not found |
|  |  | 8 | - | - | PCR false negative due to non-invasive method | Not found |
| ③ | (+, +)<br>viral presence,<br>sero-positive | 9 | + | + | various dpo | abundant |
|  |  | 10 | + | - | PPHM false negative due to low (affinity, concentration), could be seen by high sensitivity iteration of PPHM | 4 |
|  |  | 11 | - | + | PCR false negative due to viral-absence (sampling too late) | 178 |
|  |  | 12 | - | - | PCR and PPHM simultaneously false negative | Not found |
| ④ | (+, -)<br>viral presence,<br>sero-negative | 13 | + | + | high [mAb] causes PPHM false positive | Not found |
|  |  | 14 | + | - | Immunodeficiency | 2 |
|  |  | 15 | - | + | PCR false negative, high [mAb] causes PPHM false positive | Not found |
|  |  | 16 | - | - | Immunodeficiency | Not found |
| ⑤ | (-, -)<br>viral<br>clearance,<br>window period | 17 | + | + | PCR false positive, PPHM false positive | Not found |
|  |  | 18 | + | - | PCR false positive | Not found |
|  |  | 19 | - | + | PPHM false positive (other etiology) | Not found |
|  |  | 20 | - | - | IgG development lags behind virus, low dose infection | Not found |
| ⑥ | (-, +)<br>viral<br>clearance,<br>sero-positive | 21 | + | + | PCR false positive | Not found |
|  |  | 22 | + | - | PCR false positive and PPHM false negative | Not found |
|  |  | 23 | - | + | Viral clearance, sero-positive | abundant |
|  |  | 24 | - | - | PPHM false negative; Our results indicate that there is a small probability | Not found |
| ⑦ | (-, -)<br>viral<br>clearance,<br>sero-negative | 25 | + | + | PCR and PPHM false positive | Not found |
|  |  | 26 | + | - | PCR false positive | Not found |
|  |  | 27 | - | + | high [mAb] causes PPHM false positive | Not found |
|  |  | 28 | - | - | Sero-negative | Exist |

